# A WHO CNS5-Aligned Classification Framework for Population-Based Surveillance of Brain and Other Central Nervous System Tumors in the United States

**DOI:** 10.64898/2026.09.30.26364214

**Authors:** Mackenzie Price, Carol Kruchko, Daniel J. Brat, Patrick J. Cimino, Jason T. Huse, Karra A. Jones, Fernanda Michels, Tarik Tihan, Jennifer Ruhl, Quinn T. Ostrom

## Abstract

**Background:** The Central Brain Tumor Registry of the United States (CBTRUS) uses a histology grouping model based on the World Health Organization (WHO) to classify cancer registry records for clinically relevant statistical reporting.

**Methods:** CBTRUS, together with its consulting neuropathologists, reviewed and aligned its histology groupings with the 2021 WHO Classification update. Obsolete histology nomenclature along with their International Classification of Diseases for Oncology 3^rd^ edition (ICD-O-3) codes were identified, currently included histologies were reclassified as appropriate, and new codes found in 2021 WHO were incorporated. An evaluation was conducted of the frequency of histology codes affected in the reclassification process, as well as incidence and survival pre- and post-reclassification of CBTRUS Tumor Type Groupings.

**Results:** After review of the CBTRUS grouping scheme, 205 codes were reclassified and 2 new codes were incorporated. Tumor type groups most affected were those classified as glioma. Reclassification resulted in 167,038 of 491,804 (34%) cases having a grouping reclassification or reporting change, indicating that the 2021 revision of the WHO Classification has significantly impacted the collection and reporting of primary b CNS tumors.

**Conclusion:** This work demonstrates the need to be responsive to changes in classification and coding for statistics to reflect real-world clinical use. This will require collaboration from all parties to have the ability to reconcile clinical practices and surveillance requirements.

**KEY POINTS:**

- Classification changes in cancer registration pose challenges to reporting clinically-relevant population-based epidemiologic statistics for central nervous system (CNS) tumors.
- The Central Brain Tumor Registry of the United States provides an example of aligning ICD-O-3 and 2021 WHO CNS classification with cancer registry reporting.
- Implementation of ICD-O-4 and the latest WHO (CNS6) will undoubtedly be challenging to cancer registry reporting yet will ultimately improve our understanding of these tumors and their impact on a population.

**IMPORTANCE OF THE STUDY:** We performed this study to report on the Central Brain Tumor Registry of the United States (CBTRUS) tumor classification scheme which was revised to make it consistent with the 2021 WHO classification (CNS5) revision and to provide a roadmap for implementation of the recently published ICD-O 4^th^ edition. While we have previously published descriptive epidemiology for selected entities based on the CNS5 classifications, this is the first CBTRUS grouping scheme to include molecular information coded in the Brain Molecular Marker Site-Specific Data Item.

**LAY SUMMARY:** Central nervous system (CNS) tumor classification systems help doctors, researchers, and cancer registries identify and track specific types of CNS tumors. When the World Health Organization (WHO) updated its CNS tumor classification in 2021, the Central Brain Tumor Registry of the United States (CBTRUS) reviewed and updated its reporting system to match these changes. More than 200 tumor codes were reclassified, affecting nearly one-third of CNS tumor records, and most heavily impacted gliomas. These updates improve the accuracy and clinical relevance of national CNS tumor statistics. Ongoing collaboration among treating physicians, pathologists, cancer registries, and researchers will be essential as future classification updates are implemented.

## INTRODUCTION

Central nervous system (CNS) tumors represent a rare and heterogenous cancer group with over 100 distinct types. Each of these tumor types is assigned a unique histology code within the International Classification of Diseases for Oncology (ICD-O).[1] Diagnostic criteria for the classification of CNS tumors are reviewed periodically by neuropathologists and published by the World Health Organization (WHO). Most recently, the *2021 WHO Classification of Tumours of the Central Nervous System* (CNS5) was published using morphology codes from ICD-O, 3^rd^ edition to classify tumor type, behavior, and site.[2, 3]

Since its incorporation in 1992, the Central Brain Tumor Registry of the United States (CBTRUS) has become the premier research organization focused exclusively on providing high-quality statistical data on the population-based epidemiology of primary CNS tumors in the United States of America (USA).[4] CBTRUS includes data from 52 central cancer registries (CCR) (all 50 states, the District of Columbia, and Puerto Rico) provided by the Centers for Disease Control and Prevention’s (CDC) National Program of Cancer Registries (NPCR) and the National Cancer Institute’s (NCI) Surveillance, Epidemiology, and End Results (SEER) Program. The CBTRUS database is comprised of the largest tumor type-specific aggregation of population-based primary CNS tumors in the world. In collaboration with consulting neuropathologists, CBTRUS regularly updates its tumor grouping scheme to be consistent with clinical practice. This was last updated in 2021 based on the 2007 and 2016 versions of *WHO Classification of Tumours of the Central Nervous System* (CNS4 and CNS4 Revised [CNS4R] editions, respectively).[5–7]

Reporting of primary CNS tumors in the USA is mandated by state and federal law. These tumors are coded using the ICD-O, as well as other standards set by the North American Association of Central Cancer Registries (NAACCR). While the ICD-O 3^rd^ edition was updated in 2019 (ICD-O-3.2) to include many new entities introduced in CNS4R, some new molecularly-defined tumor types could not be uniquely identified using the ICD-O-3 coding scheme. To address this, CBTRUS successfully petitioned the Uniform Data Standards (UDS) Committee of NAACCR to develop the Brain Molecular Marker (BMM) Site-Specific Data Item (SSDI) to include CNS4R tumor types not identifiable through current ICD-O-3.2 codes. This variable was used nationally in 2018, with this code becoming publicly available in data in 2021.[8] This SSDI was updated to include CNS5 entities as of 2024 diagnoses, from which data will be made available in 2027.

Here, we report on the CBTRUS classification scheme revision to be consistent with CNS5 and to provide an implementation roadmap for the recently published ICD-O 4^th^ edition (ICD-O-4).[9] This is the first CBTRUS grouping scheme to include molecular information.

## METHODS

### Data source

Under a special agreement, de-identified data are directly received by CBTRUS from the CDC’s NPCR Cancer Surveillance System (NPCR-CSS) through its submission specifications mechanism[10] and are combined with data from the NCI’s SEER Program, together representing the entire U.S. population.[11] All SEER/NPCR data originate from pathology reports, radiology reports, death certificates and other data abstracted by oncology data specialists or others who adhere to the UDS for primary malignant and non-malignant CNS tumors. This dataset covers diagnosis years 1995-2022 (not all CCR included in all years due to data submission practices or NPCR/SEER data quality evaluation). Survival data from a subset of CBTRUS cases was obtained from the SEER17 for newly diagnosed cases from 2018-2022 (∼26.6% of the US population).[12]

### Reclassification process

The 2026 CBTRUS tumor grouping scheme was developed by first creating a draft document through cross-referencing five sources: (1) *CNS4R, (2) CNS5*, (3) *WHO Classification of Haematolymphoid Tumours,* (4) the ICD-O-3.2, (5) ICD-O-4, and (5) current CBTRUS classification scheme.[2, 6, 7, 9, 13, 14] Cross-referencing ensured there was complete capture of all pertinent CNS tumor types, including biomarkers utilized for diagnosis since 2018 (the first year of BMM SSDI availability). The draft grouping scheme prioritized CNS5, ICD-O-3.2, and the BMM SSDI. After development of the draft grouping scheme, CBTRUS met twice with five consulting neuropathologists, followed by three rounds of review to finalize the scheme. Large language models were used to summarize discussion and capture all review recommendations. The grouping scheme was designed to include codes not fully implemented into the UDS, as well as obsolete codes for entities not included in CNS4R or CNS5 (e.g. oligoastrocytoma, gliomatosis cerebri). These may continue to be reported despite their removal from CNS4R/CNS5, and excluding them would skew the true impact of CNS tumors. The two schemes are referred to as the *Retired CBTRUS Classification (CNS4/CNS4R*) and *New CBTRUS Classification (CNS5)* and presented as **Supplementary Table 1** and **Table 1**, respectively.

**Table 1.**
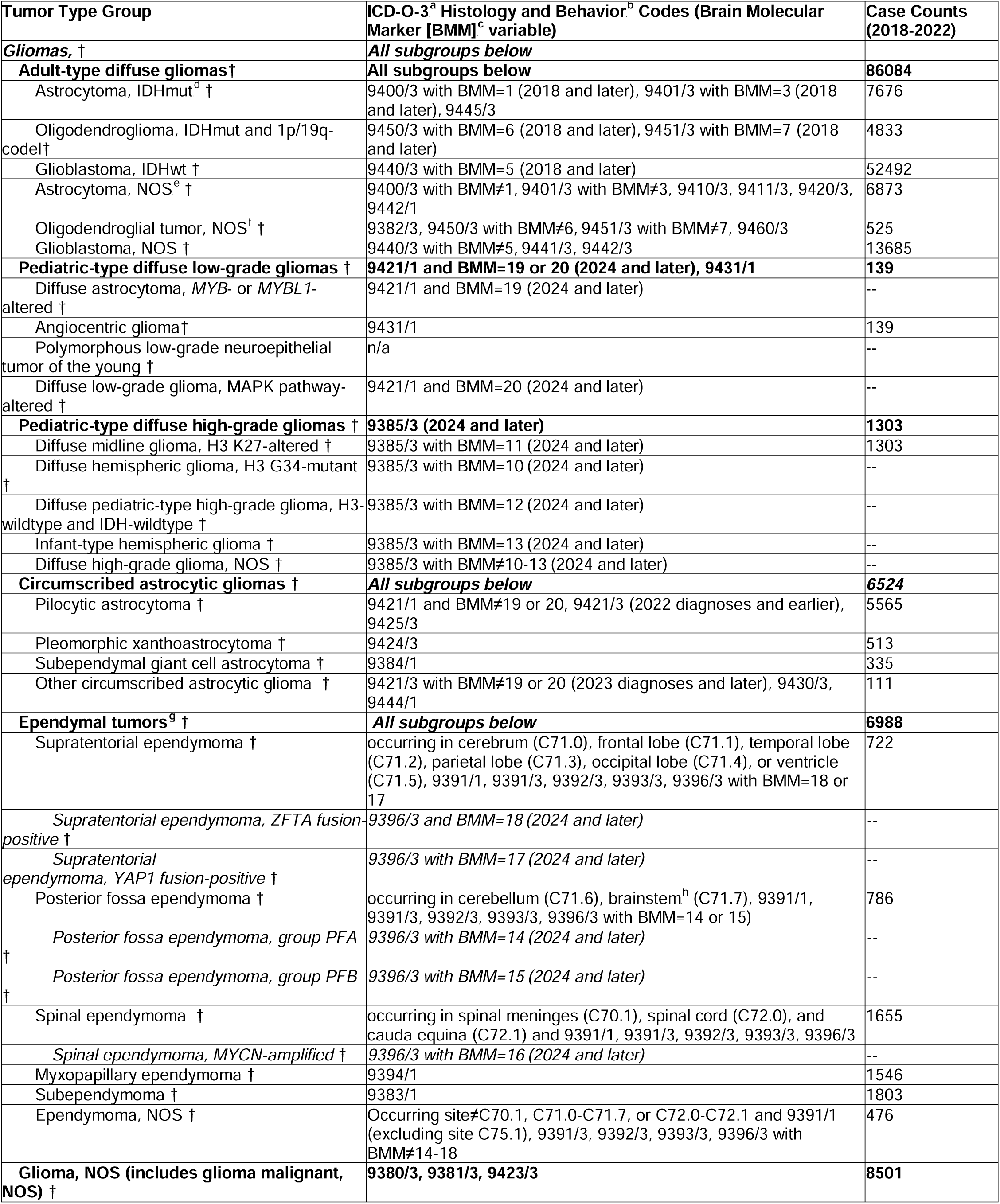
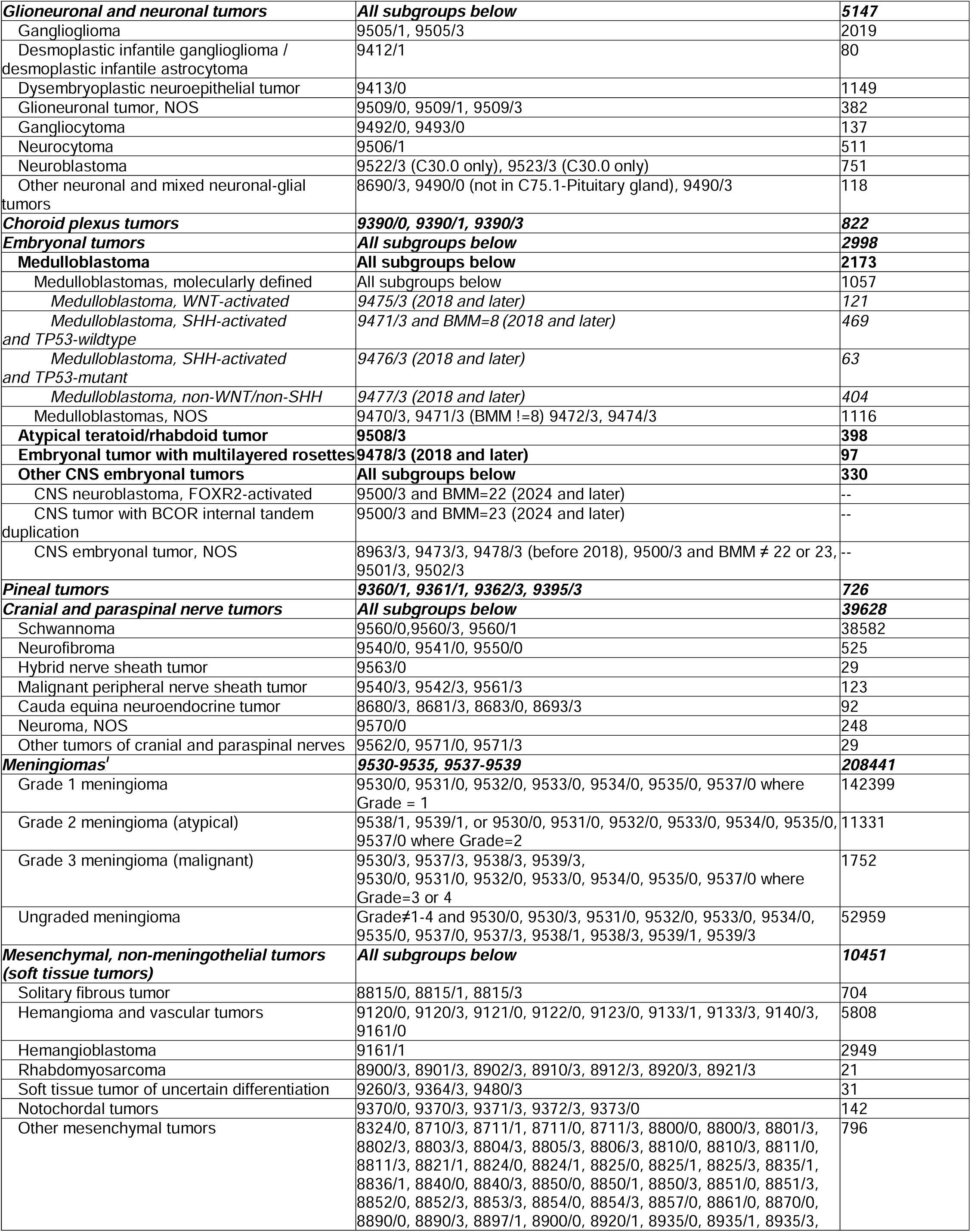
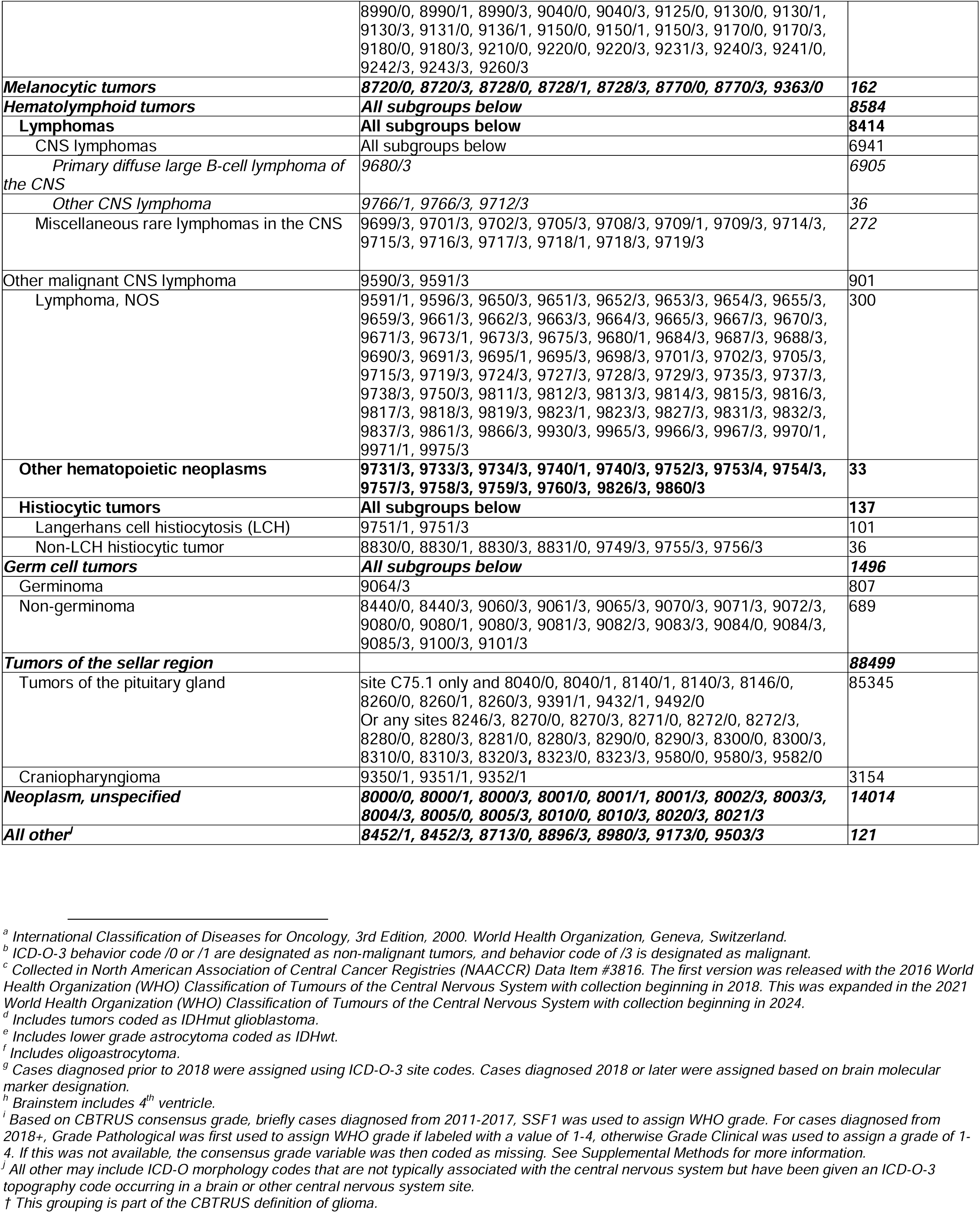
Central Brain Tumor Registry of the United States (CBTRUS) Brain and Other Central Nervous System Classification Scheme, 2026 Implementation.

Many reported tumors in the CBTRUS dataset are missing assigned grade, limiting the use of the grade variable. Due to variation in how grading variables have been used and collected over time, there are potentially conflicting grade assignments and incompatibility for pre-/post-2018 cases. CBTRUS has implemented a grade recod,[13] which was further expanded here. Briefly, in consultation with neuropathologists, we identified tumor types for which there is an obligatory grade assignment (e.g. glioblastoma is always grade 4) and used this information to recode grade assignments in conflict with histology, as well as to ‘fill in’ cases that are missing grad (See **Supplemental Methods** and **Supplementary Figure 1**).

### Statistical analysis

The CBTRUS analytic dataset was used to generate case counts for all primary CNS tumor cases from 2018-2022.[15] Counts were generated under both the *Retired* and *New Classifications*. Counts and rates are not presented when fewer than 16 cases were reported for the specific category in accordance with data use policy.

Case counts were generated by tumor type from both the *Retired* and *New Classifications* and average annual age-adjusted incidence rates (AAAIR) by tumor type from the *New Classification* for all primary malignant and non-malignant CNS tumor cases diagnosed from 1995-2022.[15] AAAIR’s were generated per 100,000 population and age-adjusted to the 2000 US Standard Population using the R package *tidyrates*.[16]

SEER data from 2018-2022 were grouped using both the *Retired* and *New Classifications*. Median survival in months, and Kaplan-Meier estimates for selected tumor types were generated with overall survival (OS). All cases were restricted to first sequence tumors and histological or radiographic confirmation. Autopsy or death certificate only cases were excluded.

Figures for were generated in R 4.5.1 using *ggplot2*, *ggsankey* and *survminer*.[17–20]

## RESULTS AND DISCUSSION

Significant modifications were made to the CBTRUS grouping scheme during the reclassification process. The *Retired Classification* consisted of 30 histology groupings (**Supplementary Table 1**). The new scheme included 14 major groups, with 66 tumor types further classified into 13 families and 11 subtypes (**Table 1**). The largest of these changes was reclassification of glioma histologies from the *Retired Classification* (*diffuse astrocytic and oligodendroglial tumors*, *other astrocytic tumors*, *ependymal tumors*, and *other gliomas*) to the *New Classification* families of *adult-type diffuse gliomas*, *pediatric-type diffuse low-grade gliomas*, *pediatric-type diffuse high-grade gliomas*, *circumscribed astrocytic gliomas*, *ependymal tumors* and *glioma, not otherwise specified (NOS)*. Overall 223 ICD-O-3.2 histology codes were reclassified (**Supplementary Table 2**).

Overall, there were 491,804 tumors from 2018-2022. Reclassification occurred for 34% (167,038) of cases. After application of the grade recode, 15,782 tumor records were re-graded (3.2%). The number of records missing grade variable was reduced from 146,310 to 136,403 (**Supplementary Figure 2**).

Major changes are described by tumor family below, and with specific tumor types definitions by ICD-O-3.2 histology, behavior, topography, and brain molecular markers codes in **Supplementary Table 4.**

### Adult-type diffuse glioma

*Adult-type diffuse glioma* was created as a new family from histologic codes previously included in the major grouping of *Diffuse Astrocytic and Oligodendroglial Tumors*. The focus was incorporation of molecularly-defined entities, rather than the prior reliance on histologically-defined entities. Separate types were retained for tumors lacking molecular classification to preserve continuity. Under the *Retired Classification* scheme, most cases are *glioblastoma* (78.6%), followed by *lower grade astrocytoma* (15.2%, including *diffuse and anaplastic astrocytoma*), and *oligodendroglioma* (6.1%, including *anaplastic oligodendroglioma*) (**Figure 1A**). Under the *New classification* scheme, the most common tumor type is *glioblastoma, IDHwt* (61%, with an additional 15.9% of cases classified as *Glioblastoma* with no molecular information). *Astrocytoma, IDHmut* (including *glioblastoma, IDHmut*) representing 8.9% of cases, while *astrocytoma, NOS* represents 8.0%. *Oligodendroglioma, IDHmut and 1p/19q-codel* represents 5.6% of cases, with <1% of cases categorized as *oligodendroglial tumor, NOS*.

**Figure 1.**
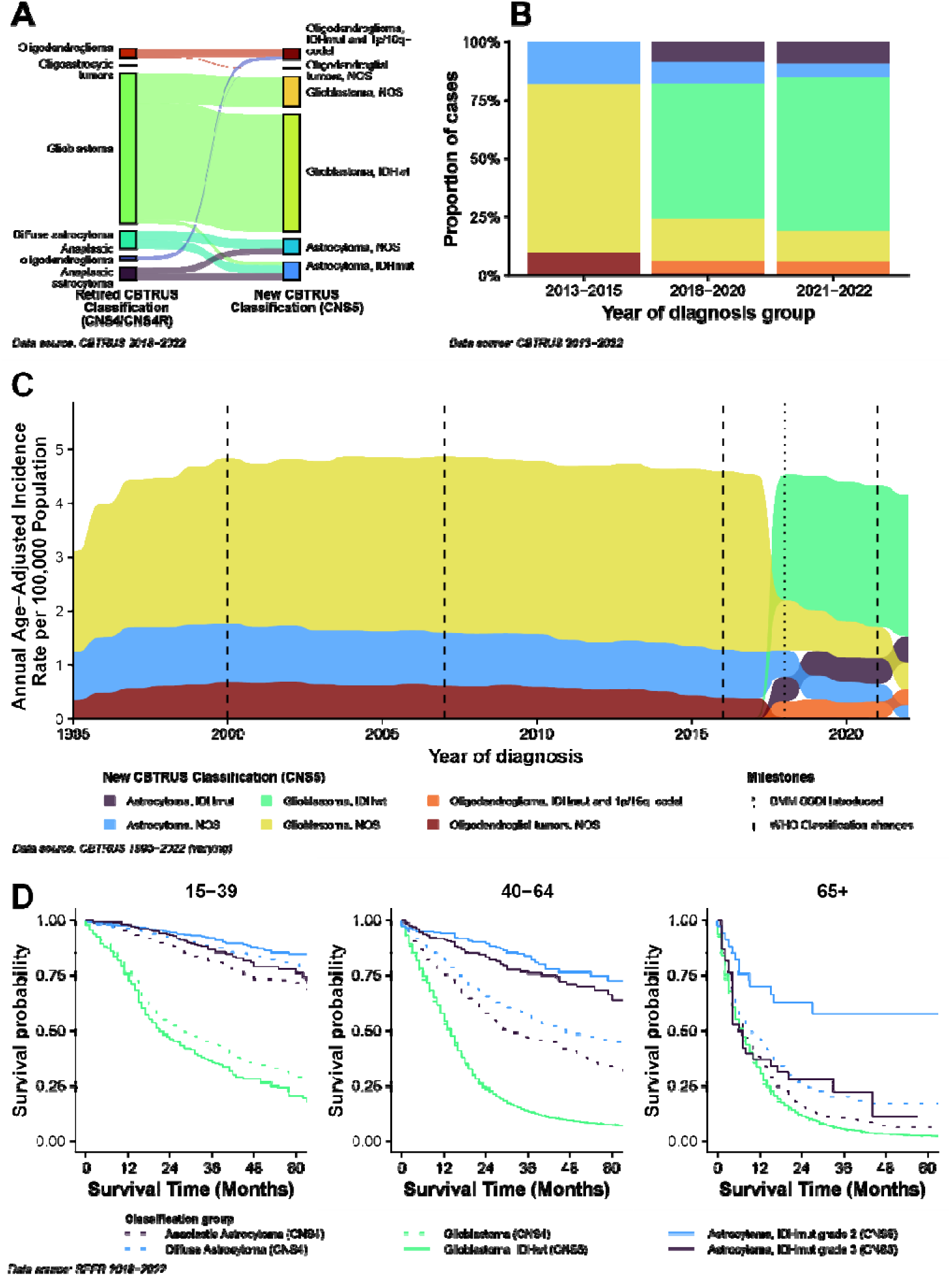
A) Changes in case classification between Retired CBTRUS Classification (CNS4/CNS4R) and New CBTRUS Classification (CNS5) scheme for adult-type diffuse glioma entities, B) proportion of adult-type diffuse glioma cases under the New CBTRUS Classification (CNS5) by year of diagnosis, C) average annual age-adjusted incidence rates of adult-type diffuse glioma entities defined by the New CBTRUS Classification (CNS5), and D) comparison of overall survival by age between new and reclassified groupings

Tumors that were classified as *Glioblastoma* under the *Retired Classification* are now categorized as *glioblastoma, IDHwt, astrocytoma, IDHmut* and *glioblastoma, NOS* (77.6%, 2.4% and 20.2%, respectively) (**Figure 2A**). Once further stratified by grade, tumors formerly categorized as *diffuse astrocytoma* are now classified as *astrocytoma, IDHmut* grade 2 (44.7%), *astrocytoma, IDHwt* grade 2 (25.2%, included in *astrocytoma, NOS*), and an additional 30% were classified as *Astrocytoma, NOS* with no molecular information. Tumors formerly categorized as *anaplastic astrocytoma* are now classified as *astrocytoma, IDHmut* grade 3 (48.1%), *astrocytoma, IDHwt* grade 3 (38.5%, included in *astrocytoma, NOS*), and an additional 13.4% were classified as *astrocytoma, NOS* with no molecular information. Proportion of agreement between the schemes varied significantly by age (**Figure 2B**), with children (ages 0-14 years old) having a greater portion of *diffuse and anaplastic astrocytoma* cases reclassified to *astrocytoma, NOS* with missing molecular information, and adolescents and young adults (AYA, ages 15-39 years old at diagnosis) more likely to have these tumors reclassified as *astrocytoma, IDHmut*.

**Figure 2.**
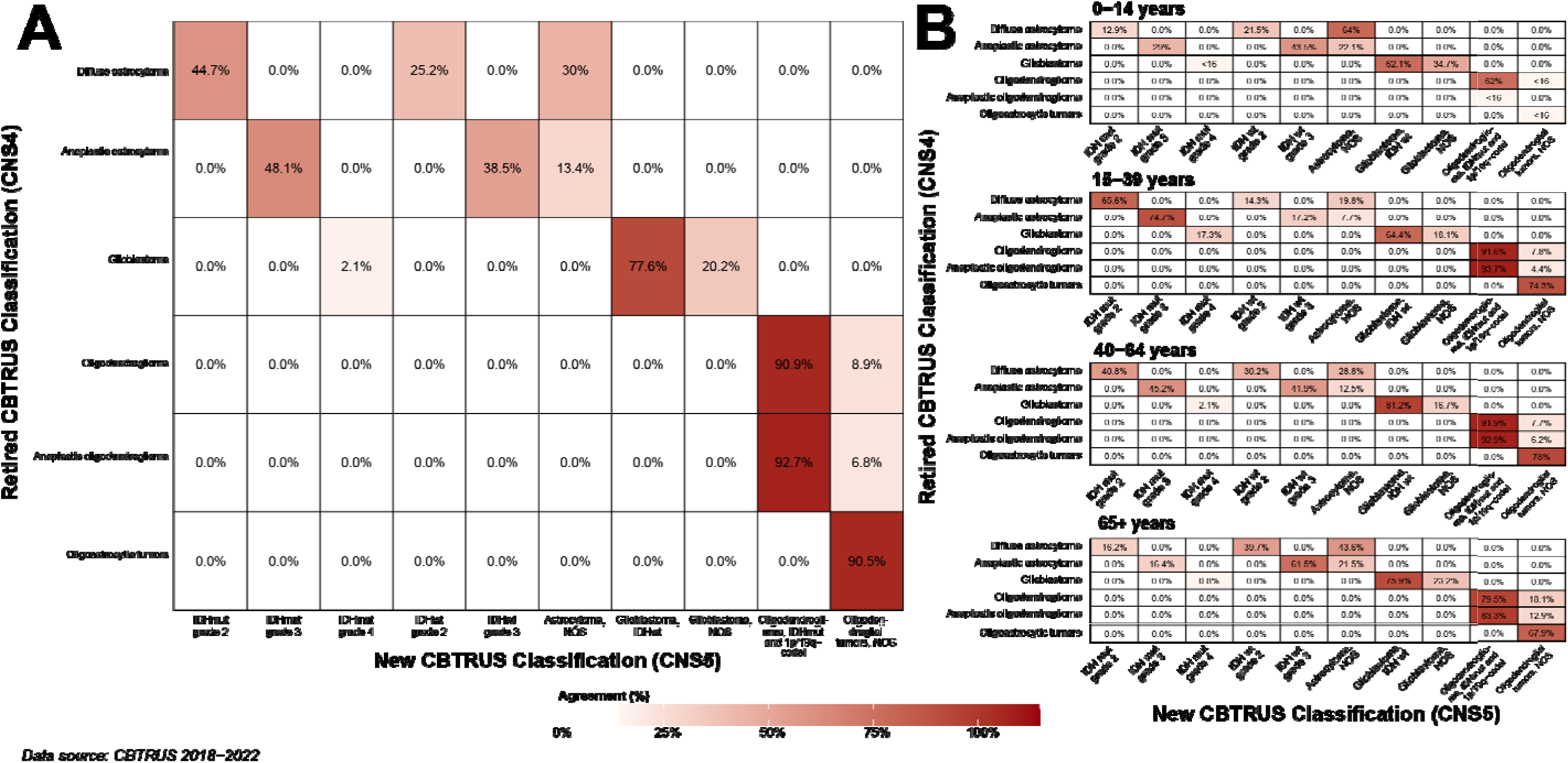
Agreement between Retired CBTRUS Classification (CNS4/CNS4R) and New CBTRUS Classification (CNS5) for selected adult-type diffuse glioma tumor types A) overall and B) by age

Not otherwise specified, or “NOS” groupings make up a significant proportion of cases within the *Adult-type diffuse glioma* family. A substantial portion of these cases are due to missing molecular data, though completeness of this data element is improving over time (**Supplementary Figure 3**).[13] One other significant limitation to incorporating complete molecular data into the cancer registry system is that the BMM SSDI is limited to specific ICD-O-3.2 codes, meaning that for tumors coded as fibrillary (9420/3) or gemistocytic (9411/3) astrocytoma (previously part of the *diffuse astrocytoma* category and now in *astrocytoma, NOS*), there is no way to code *IDH1/2* mutation, despite 70-80% of these tumors being IDHmut.[21] Both of these remain significant issues resulting in underestimation of molecular subtypes.

We calculated counts and AAAIR within time periods for *New Classification* tumor types to assess classification stability over time. During the pre-BMM era (2013-2015), all tumors were classified without molecular characterization (**Figure 1B**). The proportion of tumors classified into “NOS” groups decreased between the introductory era (2018-2020) and the two most recent years (2021-2022), because of improvements in data completeness. AAAIR of *adult-type diffuse glioma* was stable until 2018 (**Figure 1C**), when tumor type incidence changed dramatically due to introduction of the BMM SSDI.

Classification changes had substantial impacts on OS, varying by age at diagnosis (**Figure 1D**). Among AYA, where there is increased enrichment for IDH mutation in grade 4 cases, OS for *Glioblastoma* was 28 months, but OS was 21 months among the subset including only *Glioblastoma, IDHwt*. In comparison, 40-64 and 65+ age groups had minimal survival differences between the two schemes. Among older groups, where IDH mutation is less common, median OS was substantially higher for grade 2 and 3 *Astrocytoma, IDHmut* as compared to *diffuse and anaplastic astrocytoma*.

### Pediatric-type high- and low-grade diffuse glioma

The new families of *pediatric-type high-* and *low-grade diffuse glioma* were created from the *Retired Classification* major groupings of *other gliomas* and *other astrocytic tumors*, respectively (**Supplementary Figure 4A**). This incorporates the significant shift to recognize the biological distinctions between predominantly pediatric and adult diffuse gliomas. The size and complexity of these groupings will increase with the public release of the updated BMM data and implementation of ICD-O-4. With these changes, it will be possible to uniquely identify many rare tumor types within the pediatric-type high-grade glioma group, including *diffuse midline glioma, H3 K27-altered*, *diffuse hemispheric glioma, H3 G34mut*, and *diffuse pediatric-type high-grade glioma, H3wt and IDHwt*. Similarly, it will be possible to identify additional tumor types within the *pediatric-type low-grade glioma* family, including *diffuse astrocytoma, MYB- or MYBL1-altered*, and d*iffuse low-grade glioma, MAPK pathway-altered*. CBTRUS petitioned NAACCR to include a new SSDI for unique identification of the pons, midline, and other sites currently in the overall site of “Brainstem, NOS”, which will allow for most specific identification. Collection of this variable began in 2024 and will be publicly available in 2027.

### Circumscribed astrocytic gliomas

The new family was created from the prior grouping of *other astrocytic tumors*, based on the recognition that these are biologically distinct from diffuse astrocytic tumors. This family includes *pilocytic astrocytoma* (PA), *pleomorphic xanthoastrocytoma* (PXA), and *subependymal giant cell astrocytoma* (SEGA) among other rarer tumor types. Within this grouping, 85.3% are PA, while 7.8% and 5.1% are PXA and SEGA, respectively. The *New Classification* reclassifies all PA as grade 1, a significant change from the historic practice of cancer registries to reclassify these as malignant (behavior of /3).[22]

### Ependymoma

This family was reclassified by anatomic site, due to the recognition that these tumors represent biologically distinct groups associated with specific molecular alterations. While molecular data are not yet available for these tumors, location drives prognosis and clinical management. Under the *Retired Classification* scheme, all ependymoma and related tumors are classified under one *ependymal tumors* group. In the *New Classification*, the most common tumor type is *subependymoma* (25.8%), followed by *spinal ependymoma* (23.7%) and *myxopapillary ependymoma* (22.1%). *supratentorial ependymoma* and *posterior fossa ependymoma* represent 10.3% and 11.2% of cases, respectively, with 6.8% classified as *ependymoma, NOS*. The *New Classification* includes additional subtypes not currently identifiable in these data based on coming updates, including *ZFTA Fusion-Positive* and *YAP1 Fusion-Positive,* Groups *PFA and PFB,* and *MYCN-Amplified*. All *subependymoma* are reassigned grade 1 regardless of record coding.

### Embryonal tumors

Revisions to *embryonal tumors* m focused on reclassification with molecularly-defined entities in CNS5 while retaining historic categories for the purposes of surveillance. Tumor types were created for tumors missing molecular data to ensure continuity of reporting. In the *Retired Classification*, all embryonal tumors were grouped, including *Medulloblastoma, atypical/teratoid rhabdoid tumor* (ATRT), and *CNS embryonal tumor* (previously PNET). In **Figure 3A**, we show how these tumors have been subset into more precise families.

**Figure 3.**
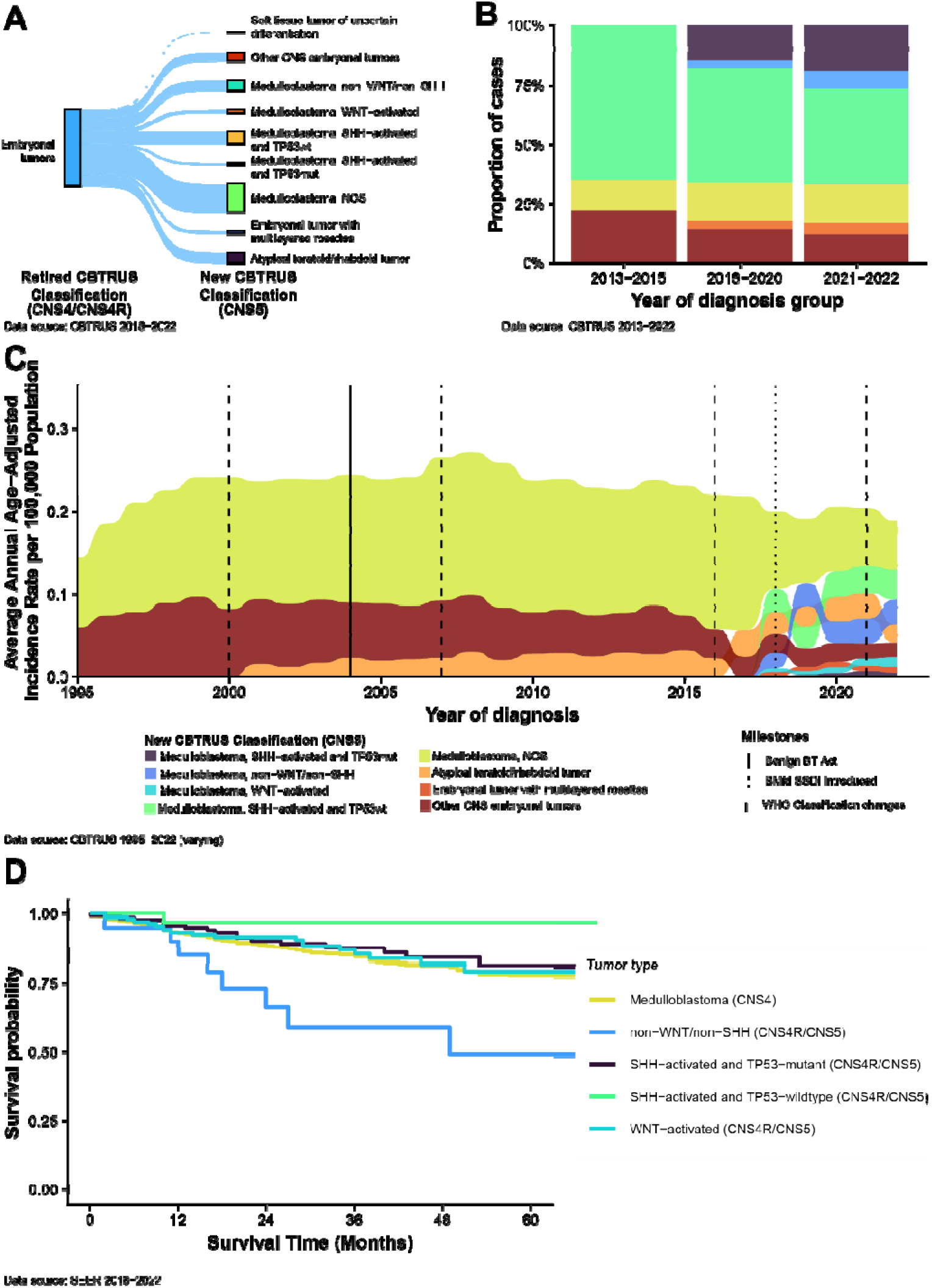
A) Changes in case classification between Retired CBTRUS Classification (CNS4/CNS4R) and New CBTRUS Classification (CNS5) scheme for embryonal tumor entities, B) proportion of embryonal tumor cases under the New CBTRUS Classification (CNS5) by year of diagnosis, C) average annual age-adjusted incidence rates of embryonal tumor entities defined by the New CBTRUS Classification (CNS5) and D) comparison of overall survival between new and reclassified groupings

The *New Classification* divides *Medulloblastoma* based on molecular information. The most common *medulloblastoma* tumor type is *medulloblastoma, NOS* (49.2%, includes cases missing molecular data). *Medulloblastoma* cases with available molecular data were further subtyped: 20.7% are SHH-activated/TP53wt, 17.8% are non-WNT/non-SHH, 5.3% are WNT-activated, and 2.8% are SHH-activated/TP53mut. Missing molecular data remains a significant problem and is likely due substantially to existing testing data results not being abstracted.

We calculated counts and AAAIR within select time periods by *New Classification* tumor types to assess its stability over time. The proportion of tumors classified into “NOS” groups decreased between the introductory era (2018-2020) and the two most recent years (2021-2022), like due to improvements in data completeness (**Figure 3B**).

Pooled AAAIR of embryonal tumors has been stable over time. Changes in definitions (e.g. the introduction of ATRT in 2001, modification of PNET classification) have had significant impacts on incidence (**Figure 3C**). After BMM SSDI introduction, incidence of tumor families varied significantly, though *Medulloblastoma, NOS* has remained the most common tumor type.

### Meningioma

The *Meningioma* major grouping was revised to focus on grade. In the *Retired Classification*, all meningiomas were pooled and frequently classified as “non-malignant” (behavior code /0 or /1, roughly equivalent to grade 1/2), and “malignant” (behavior code /3, roughly equivalent to grade 3). We now subdivide the overall category of *Meningiomas* based on the recoded grade variable, into grade 1 (68.2%), grade 2 (5.2%), grade 3 (0.8%), and ungraded (25.3%) (**Figure 4A**). Meningioma categorized as grade 4 are downgraded to grade 3, as grade 4 is not a valid grade for meningioma. Ungraded tumors are preserved to accommodate radiographically diagnosed tumors and those missing grade information.

**Figure 4.**
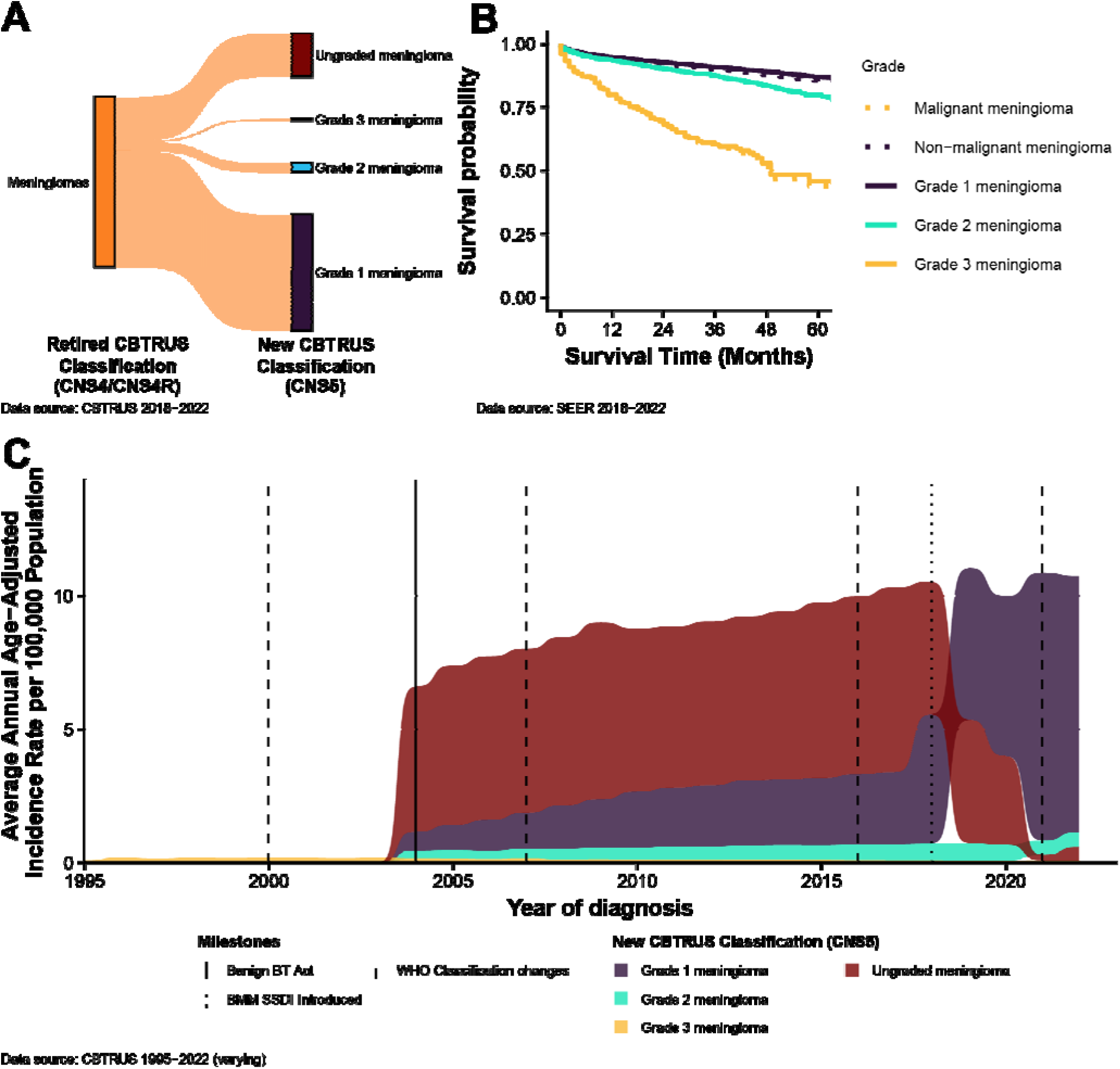
A) Changes in case classification between Retired CBTRUS Classification (CNS4/CNS4R) and New CBTRUS Classification (CNS5) scheme for meningiomas, B) comparison of overall survival between New and Retired meningioma groupings, and C) average annual age-adjusted incidence rates of meningioma entities defined by the New CBTRUS Classification (CNS5)

AAAIR have changed over time for several major reasons (**Figure 4C**). “Non-malignant” tumors were collected beginning in 2004, with significant increases in collection through 2006. Changes in grading over time after establishment of the three-tier grading system based on histologic features, followed by implementation of molecular markers in CNS5 have caused reorganization within the grade groups.[3, 6, 7, 23]

### Hematolymphoid tumors

The prior grouping of *lymphomas and hematopoietic neoplasms* was reclassified into *hematolymphoid tumors* and further separated into biologically distinct disease families. The *New Classification* divides lymphomas into *CNS lymphomas (*further subtyped to *primary diffuse large b-cell lymphoma of the CNS* [80.4%] and *other CNS lymphoma* [<1%]), *other malignant CNS lymphoma* (10.5%), *lymphoma, NOS* (4.1%), and *miscellaneous rare lymphomas in the CNS* (3.2%) (**Supplementary Figure 4B**). Codes previously included in this category were reclassified to the *histiocytic tumors* family including *Langerhans cell histiocytosis (LCH)* and *Non-LCH*. These changes reduced the size of the former *other hematopoietic neoplasms* category.

### Other tumor types

Changes to the other groupings focused on alignment with CNS5 and reducing heterogenous categories. *Nerve sheath tumors, germ cell tumors*, and *mesenchymal tumors* were reclassified into more specific tumor types (**Supplementary Figure 4C**). Several groupings and tumor types had little-to-no changes, including *choroid plexus tumors*, *tumors of the pineal region, melanocytic tumors, sellar region tumors*, and *craniopharyngioma*.

### Impact of classification changes on global CNS tumor epidemiology

The ICD-O-4 was announced in 2026 and (with input from CBTRUS) added a fifth digit to morphology codes and a fourth digit to topography codes to enable more detailed coding.[24] Current international initiatives like CONCORD and *Cancer Incidence in 5 continents*[25–27] largely use CNS4 entities for CNS tumors, and implementation of the ICD-O-4 will present significant challenges for handling historical data.

While molecular classification is now standard of care for many CNS tumor types, a small portion of individuals in high income and many in low- and middle-income countries do not receive molecular testing of medulloblastoma.[28, 29] As the ICD-O-4 is implemented, this ‘true’ missingness will likely increase due to inequities in access. This will make international comparisons increasingly difficult as a ‘two tier’ scheme of CNS tumor diagnostics continues to emerge. As targeted treatments continue to be developed for CNS tumors, this also increases inequities in access to treatment.[30, 31]

### Limitations

There were several limitations to our ability to evaluate differences in incidence and survival patterns under the *Retired* and *New Classification*s . The first is that the data used began collection January 1, 2018, predating release of CNS5. Many classification changes and molecular markers implemented in this scheme were already in use prior to 2021, so it is likely that changes could be observed prior to 2021. The second of these is that new CNS5 tumor types were not added to the BMM SSDI until 2024, and as a result are not coded in the data used.

With increasing incorporation of molecular features, CNS tumor classification becomes more precise and includes increasingly rare entities. Many of the tumor types included in this updated scheme are rare and require close attention to suppression thresholds when reporting. For these, we provide an overall ‘pooled’ family.

One inherent limitation of cancer registry data is that there is lack of central pathology review, and reported diagnosis reflects that assigned by the diagnosing pathologist based on the prevailing criteria used in that facility at that time. Some centers may adopt criteria prior to release of official changes. Academic centers may adopt procedures based on recent scientific developments for assays performed in house, while community hospitals may experience delays in adoption of changes. This may limit accurate measurement of the true population-level effect of classification changes.

## CONCLUSIONS

Changes in clinical practices significantly impact the collection and reporting of CNS tumors. Therefore, it is critical that cancer registration stays responsive to these changes to ensure that the statistics produced from these data reflect real world clinical practice. The reclassification of the CBTRUS coding system with the CNS5 demonstrates that changes in clinical practices can impact the collection and reporting of CNS tumors.

We expect that the publication of the CNS6 will incur additional updating of the CBTRUS classification scheme. CBTRUS plans to continue working with the UDS Committee to include classification changes in cancer reporting t while being mindful that any implementation will be bound by the UDS timeline.

## FUNDING

CBTRUS is honored to be included among the research awardees of the following organizations which have contributed to the analyses resulting from the CBTRUS database: the Centers for Disease Control and Prevention under Contract No. 75D30123P17556, the American Brain Tumor Association, Novocure, Jazz Pharmaceuticals, the Musella Foundation for Brain Tumor Research & Information, Inc., the National Brain Tumor Society, the Sontag Foundation, the Uncle Kory Foundation, the Josephine F. DeAngelis Memorial Fund, the Don and Diane Kane Charitable Trust, and the Zelda Dorin Tetenbaum Memorial Fund, as well as from private and in-kind donations.

## ETHICS STATEMENT

This study was conducted in accordance with the ethical principles outlined in the Declaration of Helsinki. The project was deemed exempt for review by the Duke University Health System Institutional Review Board. All data used during the study are anonymized to protect patient privacy, and no identifying information is included in the final data set.

## CONFLICTS OF INTERESTS

The authors have no relevant financial or non-financial interests to disclose.

## AUTHORSHIP

Quinn Ostrom, Mackenzie Price, and Carol Kruchko conceived of the study. The original revised classification scheme was developed by Quinn Ostrom. Daniel Brat, Tarik Tihan, Karra Jones, Patrick Cimino, Jason Huse, Mackenzie Price, Carol Kruchko, and Quinn Ostrom reviewed the scheme over multiple rounds. The first draft of the manuscript was written by Quinn Ostrom and Mackenzie Price. Analysis, and tables and figures creation was performed by Quinn Ostrom and Mackenzie Price. All authors provided feedback on the finalized scheme and commented on previous versions of the manuscript. All authors reviewed and approved the final manuscript.

## DATA AVAILABILITY

The Central Brain Tumor Registry of the United States data used in this report are not publicly available. The data used in this analysis are made available to CBTRUS through an agreement with the CDC’s National Program of Cancer Registries and the NCI’s Surveillance Epidemiology and End Results Program. Data analysis requests can be made at https://cbtrus.org. Surveillance, Epidemiology, and End Results Program data are publicly available and can be requested from http://seer.cancer.gov. Additionally, a version of these data that has not been processed using the CBTRUS edits program can be accessed through the US Cancer Statistics (https://www.cdc.gov/united-states-cancer-statistics/index.html).

## Data Availability

The Central Brain Tumor Registry of the United States data used in this report are not publicly available. The data used in this analysis are made available to CBTRUS through an agreement with the CDCs National Program of Cancer Registries and the NCIs Surveillance Epidemiology and End Results Program.

http://seer.cancer.gov

https://www.cdc.gov/united-states-cancer-statistics/index.html

https://cbtrus.org

## ACKNOWLEDGEMENTS

CBTRUS is a not-for-profit corporation affiliated with the Society for Neuro-Oncology. The CBTRUS data were provided through an agreement with the Centers for Disease Control’s (CDC) National Program of Cancer Registries. In addition, CBTRUS used data from the research data files of the National Cancer Institute’s (NCI) Surveillance, Epidemiology, and End Results Program, and the National Center for Health Statistics National Vital Statistics System. We acknowledge the oncology data specialists at the hospitals and treatment centers, the CCRs, and the staff from the NPCR and SEER programs, whose efforts to collect accurate and complete data have made this report possible. CBTRUS acknowledges and appreciates these contributions to this manuscript and to cancer surveillance in general. Contents are solely the responsibility of the authors and do not necessarily represent the official views of the CDC or the NCI. Preliminary results from this study were presented at the Brain Tumor Epidemiology Consortium and North American Association for Central Cancer Registries Annual Meetings in 2026.

## Notes

### Competing Interest Statement

The authors have declared no competing interest.

### Author Declarations

The Duke University Health System Institutional Review Board gave ethical approval for this work as a protocol exempt for review.

