## Supplementary Methods, Tables, and Figures for "A WHO CNS5-Aligned Classification Framework for Population-Based Surveillance of Brain and Other Central Nervous System Tumors in the United States"

Supplementary Table 1. Retired Central Brain Tumor Registry of the United States (CBTRUS), 2021 Brain and Other Central Nervous System Tumor Histopathology Groupings (Based on 2016 WHO Classification) ... 11
Supplementary Table 2. ICD-O-3.2 Histology codes that were reclassified ... 14
Supplementary Table 3. Definition of specific tumor types by ICD-O-3.2 histology, behavior, topography, and brain molecular markers codes ... 22
Supplementary Figure 1. Overview of recode process for specific tumor types ... 37
Supplementary Figure 2. A) Grade of glioma cases before and after grade recoding process, B) distribution of consensus grade in glioma cases, C) distribution of grade in glioma cases post-recoding process, and D) overall survival for IDHmut astrocytoma under the consensus and recoded grading schemes ... 38
Supplementary Figure 3. Completeness of the Brain Molecular Marker Variable by ICD-O-3 Code and Year of Diagnosis ... 39
Supplementary Figure 4. Changes in case classification between 2021 and 2026 scheme for a) selected glioma entities, B) CNS lymphoma, and C) other tumor types ... 40

### 1. Supplementary methods

#### 1.1. Differences in brain tumor definitions and reporting practices

Currently, NPCR, SEER, and NAACCR report primary brain and other CNS tumors differently from CBTRUS. The definition of primary brain and other CNS tumors used by these organizations in their published incidence and mortality statistics includes tumors located in the following ICD-O-3.2 sites: brain (C70.0-9), meninges (C71.0-9), and other CNS tumors (C72.0-9), but ***excludes*** lymphoma and leukemia (ICD-O-3 morphology codes 9590-9993) from all brain and other CNS sites.[1] In contrast, the CBTRUS definition includes data on all tumor morphologies located within the Consensus Conference site definition including lymphoma and other hematopoietic tumors, tumors of the pituitary gland, and olfactory tumors of the nasal cavity (see **Supplementary Methods Table A**). CBTRUS reports data on primary brain and other CNS tumors irrespective of behavior, whereas many reporting organizations may only publish rates for **malignant** brain and other CNS tumors due to the original federal mandate that focused only on **malignant** tumors, sometimes using the term "cancer" to broadly identify these tumors in their reports. These differences in definition influence the direct comparison of published rates. CBTRUS is currently engaged in ongoing collaboration with other cancer registry reporting groups, including SEER, to harmonize brain tumor reporting definitions. Ideally, these reporting differences will be minimized in the future. Reporting of hematological malignancies of the CNS—as CNS malignancies by CBTRUS and with other hematological malignancies by SEER—will likely remain.

**Supplementary Methods Table A. Central Brain Tumor Registry of the United States (CBTRUS) Brain and Other Central Nervous System Tumor Site Groupings**

| Site | ICD-O-3 Site Code |
|---|---|
| Olfactory tumors of the nasal cavity[a] | C30.0 |
| Meninges (cerebral & spinal) | C70.0-C70.9 |
| *Cerebral meninges* | *C70.0* |
| *Spinal meninges* | *C70.1* |

| | |
|---|---|
| *Meninges, NOS* | *C70.9* |
| Cerebrum | C71.0 |
| Frontal lobe of brain | C71.1 |
| Temporal lobe of brain | C71.2 |
| Parietal lobe of brain | C71.3 |
| Occipital lobe of brain | C71.4 |
| Ventricle | C71.5 |
| Cerebellum | C71.6 |
| Brainstem | C71.7 |
| Other brain | C71.8-C71.9 |
| *Overlapping lesion of brain* | *C71.8* |
| *Brain, NOS* | *C71.9* |
| Spinal cord and cauda equina | C72.0-C72.1 |
| *Spinal cord* | *C72.0* |
| *Cauda equina* | *C72.1* |
| Cranial nerves | C72.2-C72.5 |
| *Olfactory nerve* | *C72.2* |
| *Optic nerve* | *C72.3* |
| *Acoustic nerve* | *C72.4* |
| *Cranial nerve, NOS* | *C72.5* |
| Other nervous system | C72.8-C72.9 |
| *Overlapping lesion of brain and central nervous system* | *C72.8* |
| *Nervous system, NOS* | *C72.9* |
| Pituitary and craniopharyngeal duct | C75.1-C75.2 |
| *Pituitary gland* | *C75.1* |
| *Craniopharyngeal duct* | *C75.2* |
| Pineal gland | C75.3 |

a. ICD-O-3 histopathology codes 9522-9523 only.

### 1.2. Inclusion of biomarkers

The development of technologies for characterizing DNA sequence, RNA abundance as a measure of gene activity, and epigenetic alterations that affect gene expression (such as DNA methylation) have led to the discovery of several factors known as ‘biomarkers.’ Biomarkers can be used to more accurately classify these tumors than microscopic evaluation alone. With the increased recognition of the value of biomarkers for specific brain tumor types in classification, CNS4R included biomarkers for the first time with collection of biomarker data beginning in 2018. CNS5 added additional biomarkers that began collection in 2024 and will be reportable in 2027.

**Supplementary Methods Table B. Summary of Biomarkers identified for Primary Brain and Other Central Nervous System Tumors and Status by Central Cancer Registries Collection Standards**

| Gene or Marker | Tumor Type | Collected by US Cancer Registry System |
|---|---|---|
| Large deletions (missing parts of the chromosome) including the short arm of chromosome 1 (1p) and the long arm of chromosome 19 (19q) | Glioma (especially oligodendroglial tumors) [2-6] | Yes, collected as Site-specific factor 5 (2011-2017), Site-specific factor 6 (2011-2017), Site-specific disease item: Chromosome 19q Status (2018+), Site-specific disease item: Chromosome 1p Status (2018+). |
| Protein-truncating mutation in isocitrate dehydrogenase 1 (*IDH1*) or in isocitrate dehydrogenase 2 (*IDH2)* | Glioma (especially lower grade astrocytomas and oligodendroglial tumors) [2,3,7] | Yes, began in collection year 2018 (January 1), Site-specific disease item: Brain Molecular Markers (2018+). |
| Loss of function mutation in alpha thalassemia/mental retardation syndrome X-linked (*ATRX*) | Glioma (especially IDH-mutated glioma) [7-9] | No |
| Methylation of the promoter of O-6-methylguanine-DNA methyltransferase (*MGMT*) | Glioblastoma[10-12] | Yes, collected as Site-specific factor 4 (2011-2017) and site-specific disease item: MGMT (2018+). |
| Glioma-CpG island methylator phenotype (G-CIMP), Genome-wide DNA methylation | Glioblastoma[2,13] | No |
| Amplification of epidermal growth factor receptor (*EGFR*) | Glioblastoma [2,14] | No |

| | | |
|---|---|---|
| Mutation of promotor of Telomerase reverse transcriptase (*TERT*) | Glioma (oligodendroglial tumors and IDH-wildtype glioblastoma) [2,15,16] | No |
| Mutation or fusion of B-Raf (*BRAF*) | Glioma (particularly pediatric lower grade glioma)[17] | No |
| WNT-activated medulloblastoma (Group | Medulloblastoma[18-22] | Yes, began in collection year 2018 (January 1), collected via new ICD-O-3 code. |
| SHH-activated and *TP53*-mutant medulloblastoma | Medulloblastoma [18-22] | Yes, began in collection year 2018 (January 1), collected via new ICD-O-3 code. |
| SHH-activated and TP53-wildtype medulloblastoma | Medulloblastoma[18-22] | Yes, began in collection year 2018 (January 1), collected via site-specific disease item: Brain Molecular Markers (2018+). |
| non-WNT/non-SHH, Group 3 medulloblastoma subtype (also known as Group C) | Medulloblastoma[18-22] | Yes, began in collection year 2018 (January 1), combined with group 4 and collected via new ICD-O-3 code. |
| non-WNT/non-SHH, Group 4 medulloblastoma subtype (also known as Group D) | Medulloblastoma[18-22] | Yes, began in collection year 2018 (January 1), combined with group 3 and collected via new ICD-O-3 code. |
| C19MC amplification and presence of multilayered rosettes | Embryonal tumor[23,24] | Yes, began in collection year 2018 (January 1), collected via Site-specific disease item: Brain Molecular Markers (2018+). |

As of 2011, SEER registries began collecting information on three validated biomarkers for primary brain and other CNS tumors as Site-Specific Factors (SSF): promoter methylation status of *O-6-Methylguanine-DNA Methyltransferase* (*MGMT)*, codeletion of 1p and 19q.[25] Starting with diagnosis year 2018, the broad US cancer registry system began collecting information on multiple brain and other CNS markers, including isocitrate dehydrogenase 1/2 (*IDH1/2*) mutation, 1p/19q codeletion, medulloblastoma molecular subtypes, and all biomarkers found in CNS4R using the variable Brain Molecular Markers Site-Specific Data Item (please see **Supplementary Methods Table C**). Additional molecularly-defined entities from CNS4R were added using their new ICD-O-3.2 codes for which collection also began in 2018. However, implementing the collection of these markers in cancer registration is multi-faceted and includes software updates, as well as an ongoing educational and training component.

These data were available to CBTRUS for the first time with the 2021 NPCR and SEER data releases. As such, these data are for the 2018-2022 diagnosis years only. CBTRUS evaluated the completeness of these markers in their first year (2018) of collection (please see Iorgulescu, et al.[26]). The collection of new molecularly defined entities, introduced in CNS5, began with diagnosis year starting January 1, 2024 for the first time and will be available for analysis by CBTRUS in reporting year 2027.

**Supplementary Methods Table C. Coding Definitions for the Brain Molecular Markers Data Item (NAACCR Data Item #3816)**

| Code | Description |
|---|---|
| 01 | Diffuse astrocytoma, IDH-mutant (9400/3) |
| 02 | Diffuse astrocytoma, IDH-wildtype (9400/3) |
| 03 | Anaplastic astrocytoma, IDH-mutant (9401/3) |
| 04 | Anaplastic astrocytoma, IDH-wildtype (9401/3) |
| 05 | Glioblastoma, IDH-wildtype (9440/3) |
| 06 | Oligodendroglioma, IDH-mutant and 1 p/19q co-deleted (9450/3) |
| 07 | Anaplastic oligodendroglioma, IDH-mutant and 1p/19q co-deleted (9451/3) |
| 08 | Medulloblastoma, *SHH*-activated and *TP53*-wildtype (9471/3) |
| 09 | Embryonal tumor with multilayered rosettes, C19MC-altered (9478/3) |

| | |
|---|---|
| 10 | Diffuse hemispheric glioma, H3-G34 mutant (9385/3) * |
| 11 | Diffuse midline glioma, H3 K27-altered (9385/3) |
| 12 | Diffuse pediatric-type high-grade glioma, H3-wildtype and IDH-wildtype (9385/3)* |
| 13 | Infant-type hemispheric glioma (9385/3)* |
| 14 | Posterior fossa group A (PFA) ependymoma (9396/3)* |
| 15 | Posterior fossa group B (PFB) ependymoma (9396/3)* |
| 16 | Spinal ependymoma, MYCN-amplified (9396/3)* |
| 17 | Supratentorial ependymoma, YAP1 fusion-positive (9396/3)* |
| 18 | Supratentorial ependymoma, ZFTA fusion-positive (9396/3) |
| 19 | Diffuse astrocytoma, MYB- or MYBL1-altered (9421/1) * |
| 20 | Diffuse low-grade glioma, MAPK pathway-altered (9421/1) * |
| 21 | Astroblastoma, MN1-altered (9430/3) |
| 22 | CNS neuroblastoma, FOXR2-activated (9500/3)* |
| 23 | CNS tumor with BCOR internal tandem duplication (9500/3) * |
| 85 | Not applicable: Morphology code not 9385/3, 9396/3, 9400/3, 9401/3, 9430/3, 9440/3, 9450/3, 9451/3, 9471/3, 9478/3, 9421/1, 9430/3, 9500/3 |
| 86 | Benign or borderline tumor (Excludes: 9421/1 (codes 19-20)) |
| 87 | Test ordered, results not in chart |
| 88 | Not applicable: Information not collected for this case (If this item is required by your standard setter, use of code 88 will result in an edit error.) |
| 99 | Not documented in patient record; No microscopic confirmation; Brain molecular markers not assessed or unknown if assessed |

a. This data item applies only to ICD-O-3 morphology codes: 9385/3, 9396/3, 9400/3, 9401/3, 9430/3, 9440/3, 9450/3, 9451/3, 9471/3, 9478/3, 9421/1, 9430/3, and 9500/3. If a morphology code is not included in this list, code 85 was assigned. Only one code is applicable for each tumor.
Abbreviations: NAACCR, North American Association of Central Cancer Registries; CNS, central nervous system.

### 1.3. Recode of ICD-O-3.2 behavior code for invalid codes

To reconcile histologic behavior codes that did not correspond to a valid entry in the ICD-O-3.2 Histology and Behavior code lists (as provided by SEER at https://seer.cancer.gov/icd-o-3/), we implemented an application of an iterative recoding procedure. Each unique morphology code present in the CBTRUS dataset was checked against both reference lists (ICD-O-3.2 and CNS5). Codes already present in either reference list were left unmodified. For codes absent from both reference lists, an initial behavior-code substitution was attempted: behavior code 1 (borderline) was recoded to 0 (benign), 3 (malignant) was recoded to 1, and 0 was recoded to 3. The histology code was then re-concatenated with the newly assigned behavior code to form an updated merged code. This updated code was then checked against the reference lists. If it still did not match either list, alternate substitution schemes were applied to the original behavior codes (1→3, 3→0, 0→1), and the new code was regenerated accordingly. If the first substitution produced a code that matched a reference list, no further recoding was performed for that code. This process was repeated across all unique unmatched codes, producing a revised behavior variable that maximized concordance with standard morphology coding conventions. A total of 6,113 cases had behavior variable recoded. See **Supplementary Methods Table D** for a summary of recoded codes.

#### Supplementary Methods Table D. Invalid codes, recoded codes, and count of affected cases

| Existing ICD-O-3.2 code | Recoded ICD-O-3.2 code | Case Counts (2018-2022) | Name |
|---|---|---|---|
| 8004/1 | 8004/3 | <16 | Malignant tumor, spindle cell type |
| 8271/3 | 8271/0 | <16 | Prolactinoma |
| 8320/0 | 8320/3 | <16 | Granular cell carcinoma |
| 8680/0 | 8680/1 | 59 | Paraganglioma, NOS |
| 8690/0 | 8690/3 | <16 | Middle ear paraganglioma |
| 8690/1 | 8690/3 | <16 | Middle ear paraganglioma |
| 8693/0 | 8693/3 | <16 | Extra-adrenal paraganglioma, malignant |

| | | | |
|---|---|---|---|
| 8693/1 | 8693/3 | 34 | Extra-adrenal paraganglioma, malignant |
| 8713/1 | 8713/0 | <16 | Glomangiomyoma |
| 8831/3 | 8831/0 | <16 | Histiocytoma, NOS |
| 8836/0 | 8836/1 | <16 | Angiomatoid fibrous histiocytoma |
| 9064/0 | 9064/3 | <16 | Germinoma |
| 9120/1 | 9120/0 | <16 | Hemangioma, NOS |
| 9161/3 | 9161/1 | 17 | Hemangioblastoma |
| 9173/3 | 9173/0 | <16 | Cystic lymphangioma |
| 9241/3 | 9241/0 | <16 | Chondromyxoid fibroma |
| 9350/3 | 9350/1 | <16 | Craniopharyngioma |
| 9352/0 | 9352/1 | <16 | Papillary craniopharyngioma |
| 9361/3 | 9361/1 | <16 | Pineocytoma |
| 9362/0 | 9362/3 | <16 | Pineoblastoma |
| 9363/3 | 9363/0 | <16 | Melanotic neuroectodermal tumor |
| 9381/0 | 9381/3 | <16 | Gliomatosis cerebri |
| 9383/3 | 9383/1 | <16 | Subependymoma |
| 9384/3 | 9384/1 | <16 | Supependymal giant cell astrocytoma |
| 9394/3 | 9394/1 | 17 | Myxopapillary ependymoma |
| 9412/3 | 9412/1 | <16 | Desmoplastic infantile astrocytoma |
| 9413/1 | 9413/0 | <16 | Dysembryoplastic neuroepithelial tumor |
| 9413/3 | 9413/0 | <16 | Dysembryoplastic neuroepithelial tumor |
| 9421/3 | 9421/1 | 4626 | Pilocytic astrocytoma |
| 9424/0 | 9424/3 | <16 | Pleomorphic xanthoastrocytoma |
| 9431/3 | 9431/1 | <16 | Angiocentric glioma |
| 9451/1 | 9451/3 | <16 | Oligodendroglioma, anaplastic |
| 9490/1 | 9490/0 | <16 | Ganglioneuroma |
| 9506/0 | 9506/1 | <16 | Centrol neurocytoma |
| 9506/3 | 9506/1 | <16 | Centrol neurocytoma |
| 9531/1 | 9531/0 | 18 | Meningothelial meningioma |
| 9531/3 | 9531/0 | <16 | Meningothelial meningioma |
| 9532/1 | 9532/0 | <16 | Fibrous meningioma |
| 9532/3 | 9532/0 | <16 | Fibrous meningioma |
| 9533/1 | 9533/0 | <16 | Psammomatous meningioma |
| 9533/3 | 9533/0 | <16 | Psammomatous meningioma |
| 9534/1 | 9534/0 | <16 | Angiomatous meningioma |
| 9535/1 | 9535/0 | <16 | Hemangioblastic meningioma |
| 9535/3 | 9535/0 | <16 | Hemangioblastic meningioma |
| 9537/1 | 9537/0 | <16 | Transitional meningioma |
| 9537/3 | 9537/0 | <16 | Transitional meningioma |
| 9541/1 | 9541/0 | <16 | Melanotic neurofibroma |
| 9561/0 | 9561/3 | <16 | MPNST with rhabdomyoblastic differentiation |
| 9570/1 | 9570/0 | <16 | Neuroma, NOS |
| 9680/0 | 9680/1 | <16 | EBV positive mucocutaneous ulcer |

### 1.4. Classification by CNS WHO grade

Primary brain and other CNS tumors generally do not disseminate outside of the CNS (with some exceptions, such as medulloblastoma), and therefore most of these tumor types are not staged according to the American Joint Commission of Cancer (AJCC) schema like other types of solid cancers. Instead, they are classified according to the *WHO Classification of Tumours of the Central Nervous System* which assigns a CNS WHO grade (grade I through grade IV assigned in CNS4R and earlier, with grades 1 through 4 in the CNS5) based on predicted clinical behavior. The WHO classification scheme was first released in 2000, and although it was updated in 2007 and 2016,[27] these updated schema were not fully implemented by US CCRs until diagnosis year 2019 or reporting year 2022. Updates made in 2007 and 2016 may affect diagnostic practices used in characterization of individual tumors included in this report. Significant changes were made to grading nomenclature and criteria in the CNS5 and this may affect cases diagnosed in 2021 and later that are included in this manuscript.

The WHO grading assignments have been recorded by cancer registrars as Collaborative Stage Site-Specific Factor 1 - WHO Grade Classification (SSF1) as directed in the AJCC, 8th Edition, Chapter 72 on Brain and Spinal Cord[28] (cases diagnosed from 2011-2017) (see **Supplementary Methods Table D** for an overview of this coding scheme), but are now recorded using Site-Specific Data Items (SSDI) Grade Pathological (cases diagnosed 2018 or later that have been resected and for which no neoadjuvant therapy was administered, NAACCR #3844), and SSDI Grade Clinical (cases diagnosed 2018 or later before any treatment has been received, NAACCR #3843) (see **Supplementary Methods Table E** for an overview of coding for these SSDI coding variables).[29] Site-specific factor variables were a required component of cancer registry data collection for brain and other CNS tumors beginning in 2004 for SEER registries and beginning in 2011 for NPCR registries, and these factors were collected through 2017 at which point they were replaced with SSDI. Completeness of these variables have improved significantly over time.[25,30]

**Supplementary Methods Table D. Overview of CS SSF1 coding of WHO**

| Code | Description |
|---|---|
| 010 | Grade I |
| 020 | Grade II |
| 030 | Grade III |
| 040 | Grade IV |
| 988 | Not applicable: Information not collected for this case |
| 998 | No histologic examination of primary site |
| 999 | Not documented in medical record; Unknown; WHO grade not stated |

**Supplementary Methods Table E. Overview of coding used for "Grade Pathological" and "Grade Clinical" variables**

| Code | Description |
|---|---|
| 1 | WHO Grade I: Circumscribed tumors of low proliferative potential associated with the possibility of cure following resection |
| 2 | WHO Grade II: Infiltrative tumors with low proliferative potential with increased risk of recurrence |
| 3 | WHO Grade III: Tumors with histologic evidence of malignancy, including nuclear atypia and mitotic activity, associated with an aggressive clinical course |
| 4 | WHO Grade IV: Tumors that are cytologically malignant, mitotically active, and associated with rapid clinical progression and potential for dissemination |
| L† | Stated as "low-grade" NOS |
| H† | Stated as "high-grade" NOS |
| A† | Well differentiated |
| B† | Moderately differentiated |
| C† | Poorly differentiated |
| D† | Undifferentiated, anaplastic |
| 9 | Grade cannot be assessed (GX); Unknown |

† Not utilized in CBTRUS grading system, considered to be missing grade information

Completeness of WHO grade is defined as having a value equal to WHO grade 1, 2, 3, or 4. Cases where WHO grade is marked as 'not applicable' or 'not documented' are considered incomplete. It is not possible to conclusively determine WHO grade, which is based on the appearance of tumor cells, when a tumor is radiographically-confirmed only. Some tumor types (including tumors of the pituitary gland and lymphomas) are often not assigned a WHO grade. This information may also be assigned but not included in the pathology report.

#### 1.5. Assignment of histology-based grading

Many reported tumor types in the CBTRUS analytical dataset are missing assigned WHO grade, limiting the use of the grade variable for reporting. Due to variation in how grading variables have been used and collected over time, there are also potentially conflicting grade assignments as well as incompatibility for pre- and post-2018 cases.

To harmonize these grade variables, we created a consensus grade variable. For cases diagnosed from 2011–2017, SSF1 was used to assign WHO grade. For cases diagnosed from 2018+, Grade Pathological was first used to assign WHO grade if labeled with a value of 1-4. If Grade Pathological could not be used to assign a grade, Grade Clinical was used to assign a grade of 1-4. If this was not available, the consensus grade variable was then coded as missing.

In consultation with the CBTRUS Consulting Neuropathologists (Drs. Brat, Cimino, Huse, Jones, and Tihan), we developed a grade re-code for selected glioma subtypes for which the brain molecular markers SSDI is applicable and selected other tumor types such as meningioma. (see **Supplementary Figure 1** below)

Briefly, tumors with an ICD-O-3 code of glioblastoma (9440/3, 9441/3, 9442/3, or 9445/3) were recoded to have a grade of 4, regardless of consensus grade value. Diffuse astrocytoma (ICD-O-3 9400/3) were obligate grade 2 tumors by the CNS4R WHO classification, and therefore these were recoded to have a grade of 2 irrespective of consensus grade. As anaplastic astrocytoma (ICD-O-3 (9401/3) and anaplastic oligodendroglioma tumors (ICD-O-3 codes of 9451/3 or 9460/3) are obligate grade 3 tumors based on CNS4R, these were recoded to have a grade of 3 regardless of originally coded grade. Irrespective of reported behavior pilocytic astrocytoma, subependymoma, and subependymal giant cell astrocytoma are coded as grade 1,myxopapillary ependymoma and fibrillary astrocytoma are given a grade 2, and medulloblastoma, ATRT, and embryonal tumors with multilayered rosettes are grade 4. Meningioma with behavior /0 that are assigned grade 4 were given a grade 3. Meningioma with behavior /1 are assigned grade 2 and meningioma with behavior /3 are assigned grade 3. For all other tumor types, consensus grade was used as the grade for analysis.

### 2. Supplementary results

#### Supplementary Table 1. Retired Central Brain Tumor Registry of the United States (CBTRUS), 2021 Brain and Other Central Nervous System Tumor Histopathology Groupings (Based on 2016 WHO Classification)

| Histopathology | ICD-O-3[i] Histopathology Codes[ii] | ICD-O-3[i] Histopathology and Behavior Code | |
|---|---|---|---|
| | | Malignant | Non-Malignant |
| *Diffuse Astrocytic and Oligodendroglial Tumors* | | | |
| Diffuse astrocytoma* | 9381, 9400, 9410, 9411, 9420, 9442/1 | 9381/3, 9400/3, 9410/3, 9411/3, 9420/3 | 9442/1 |
| Anaplastic astrocytoma* | 9401 | 9401/3 | *None* |
| Glioblastoma* | 9440, 9441, 9442/3, 9445[iii] | 9440/3, 9441/3, 9442/3, 9445/3 | *None* |
| Oligodendroglioma* | 9450 | 9450/3 | *None* |
| Anaplastic oligodendroglioma* | 9451, 9460 | 9451/3, 9460/3 | *None* |
| Oligoastrocytic tumors* | 9382 | 9382/3 | *None* |
| *Other Astrocytic Tumors* | | | |
| Pilocytic astrocytoma* | 9421, 9425[iii] | 9421/1[iv], 9421/3,9425/3 | *None* |
| Unique astrocytoma variants* | 9384, 9424, 9431[iii] | 9424/3 | 9384/1, 9431/1 |
| *Ependymal tumors** | 9383, 9391 (excluding site C75.1 for behavior /1), 9392- 9394, 9396[iii] | 9391/3, 9392/3, 9393/3, 9396/3 | 9383/1, 9391/1 (excluding site C75.1), 9394/1 |
| *Other Gliomas* | | | |
| Glioma malignant, NOS* | 9380, 9385[iii] | 9380/3, 9385/3 | *None* |
| Other neuroepithelial tumors* | 9423, 9430, 9444 | 9423/3, 9430/3 | 9444/1 |
| *Neuronal and Mixed Neuronal-Glial Tumors** | 8680, 8681, 8690, 8693, 9412, 9413, 9490, 9492 (excluding site C75.1), 9493, 9505, 9506, 9509[iii], 9522 (site C30.0 only), 9523 (site C30.0 only) | 8680/3, 8693/3, 9490/3, 9505/3, 9509/3, 9522/3 (site C30.0 only), 9523/3 (site C30.0 only) | 8680/0,1, 8681/1, 8690/1, 8693/1, 9412/1, 9413/0, 9490/0, 9492/0 (excluding site C75.1), 9493/0, 9505/0,1, 9506/1, 9509/1 |
| *Choroid Plexus Tumors* | 9390 | 9390/3 | 9390/0,1 |
| *Tumors of The Pineal Region* | 9360, 9361, 9362, 9395[iii] | 9362/3, 9395/3 | 9360/1, 9361/1 |
| *Embryonal Tumors* | 8963, 9364, 9470-9478[c], 9480, 9500, 9501/3, 9502/3, 9508 | 8963/3, 9364/3, 9470/3, 9471/3, 9472/3, 9473/3, 9474/3, 9475/3, 9476/3, 9477/3, 9478/3, 9480/3, 9500/3, 9501/3, 9502/3, 9508/3 | *None* |
| Medulloblastoma | 9470-9472,9474-9478 | 9470/3, 9471/3, 9472/3, 9474/3, 9475/3, 9476/3, 9477/3, 9478/3, | *None* |
| Atypical teratoid/rhabdoid tumor | 9508 | 9508/3 | *None* |

| | | | |
|---|---|---|---|
| Other embryonal tumors[v] | 8963, 9364, 9473, 9480, 9500, 9501, 9502 | 8963/3, 9364/3, 9473/3, 9480/3, 9500/3, 9501/3, 9502/3 | *None* |
| *Tumors of Cranial and Paraspinal Nerves* | | | |
| Nerve sheath tumors | 9540, 9541, 9550, 9560, 9561, 9570, 9571 | 9540/3, 9560/3, 9561/3, 9571/3 | 9540/0,1, 9541/0, 9550/0, 9560/0,1, 9570/0, 9571/0 |
| Other tumors of cranial and paraspinal nerves | 9562, 9563 | *None* | 9562/0, 9563/0 |
| *Tumors of Meninges* | | | |
| Meningioma | 9530-9535, 9537-9539 | 9530/3, 9537/3, 9538/3, 9539/3 | 9530/0,1, 9531/0, 9532/0, 9533/0, 9534/0, 9535/0, 9537/0, 9538/1, 9539/1 |
| Mesenchymal tumors | 8324, 8710, 8711, 8800-8806, 8810, 8811, 8815, 8821, 8824, 8825, 8830, 8831, 8835, 8836, 8840, 8850-8854, 8857, 8861, 8870, 8880, 8890, 8897, 8900-8902, 8910, 8912, 8920, 8921, 8935, 8990, 9040, 9120, 9125, 9130, 9131, 9136, 9150, 9161, 9170, 9180, 9210, 9220, 9231, 9240, 9241, 9243, 9260, 9370-9373 | 8710/3, 8711/3, 8800/3, 8801/3, 8802/3, 8803/3, 8804/3, 8805/3, 8806/3, 8810/3, 8811/3, 8815/3[iii], 8825/3, 8830/3, 8840/3, 8850/3, 8851/3, 8852/3, 8853/3, 8854/3, 8857/3, 8890/3, 8900/3, 8901/3, 8902/3, 8910/3, 8912/3, 8920/3, 8921/3, 8935/3, 8990/3, 9040/3, 9120/3, 9130/3, 9150/3, 9170/3, 9180/3, 9220/3, 9231/3, 9240/3, 9243/3, 9260/3, 9370/3, 9371/3, 9372/3 | 8324/0, 8711/0, 8800/0, 8810/0, 8811/0, 8815/0,1[iii], 8821/1, 8824/0,1, 8825/0,1, 8830/0,1, 8831/0, 8835/1, 8836/1, 8840/0, 8850/0,1, 8851/0, 8852/0, 8854/0, 8857/0, 8861/0, 8870/0, 8880/0, 8890/0,1, 8897/1, 8900/0, 8920/1, 8935/0,1, 8990/0,1, 9040/0, 9120/0, 9125/0, 9130/0,1, 9131/0, 9136/1, 9150/0,1, 9161/0,1, 9170/0, 9180/0, 9210/0, 9220/0, 9241/0, 9373/0 |
| Primary melanocytic lesions | 8720, 8728, 8770 | 8720/3, 8728/3, 8770/3 | 8728/0,1, 8770/0 |
| *Lymphomas and Hematopoietic Neoplasms* | | | |
| Lymphoma | 9590, 9591, 9596, 9650-9655, 9659, 9661-9665, 9667, 9670, 9671, 9673, 9675, 9680, 9684, 9687, 9688, 9690, 9691, 9695, 9698, 9699, 9701, 9702, 9705, 9712, 9714, 9715, 9719, 9724, 9727-9729, 9735, 9737, 9738, 9750, 9751, 9755, 9756, 9811-9819, 9823, 9826, 9827, 9831, 9832, 9837, 9861, 9866, 9930, 9965, 9966, 9967, 9970, 9971, 9975 | 9590/3, 9591/3, 9596/3, 9650/3, 9651/3, 9652/3, 9653/3, 9654/3, 9655/3, 9659/3, 9661/3, 9662/3, 9663/3, 9664/3, 9665/3, 9667/3, 9670/3, 9671/3, 9673/3, 9675/3, 9680/3, 9684/3, 9687/3, 9688/3, 9690/3, 9691/3, 9695/3, 9698/3, 9699/3, 9701/3, 9702/3, 9705/3, 9712/3, 9714/3, 9715/3, 9719/3, 9724/3, 9727/3, 9728/3, 9729/3, 9735/3, 9737/3, 9738/3, 9750/3, 9751/3, 9755/3, 9756/3, 9811/3, 9812/3, 9813/3, 9814/3, 9815/3, 9816/3, 9817/3, 9818/3, 9819/3, 9823/3, 9826/3, 9827/3, 9831/3, 9832/3, 9837/3, 9861/3, 9866/3, 9930/3, 9965/3, 9966/3, 9967/3, 9971/3, 9975/3 | 9750/1, 9751/1, 9970/1 |
| Other hematopoietic neoplasms | 9731, 9733, 9734, 9740, 9741, 9749, 9752-9754, 9757-9760, 9766, 9860, | 9731/3, 9733/3, 9734/3, 9740/3, 9741/3, 9749/3, 9753/3, 9754/3, 9757/3, 9758/3, 9759/3, 9760/3, 9766/3, 9860/3, | 9740/1, 9752/1, 9753/1, 9766/1 |
| *Germ Cell Tumors* | 8440, 9060, 9061, 9064, 9065, 9070-9072, 9080-9083, 9084/3, 9085, 9100, 9101 | 8440/3, 9060/3, 9061/3, 9064/3, 9065/3, 9070/3, 9071/3, 9072/3, 9080/3, 9081/3, 9082/3, 9083/3, 9084/3, 9085/3, 9100/3, 9101/3 | 8440/0, 9080/0,1 |

| | | | |
|---|---|---|---|
| *Tumors of Sellar Region* | | | |
| Tumors of the pituitary | 8040 (site C75.1 only), 8140 (site C75.1 only), 8146 (site C75.1 only), 8246, 8260 (site C75.1 only), 8270-8272, 8280, 8281, 8290, 8300, 8310, 8323, 9391/1 (site C75.1 only), 9432[iii] (site C75.1 only), 9492 (site C75.1 only), 9580, 9582 | 8140/3, 8246/3, 8260/3, 8270/3, 8272/3, 8280/3, 8281/3, 8290/3, 8300/3, 8310/3, 8323/3, 9580/3 | 8040/0,1, 8140/0,1, 8146/0, 8260/0, 8270/0, 8271/0, 8272/0, 8280/0, 8281/0, 8290/0, 8300/0, 8310/0, 8323/0, 9391/1 (site C75.1 only), 9432/1, 9492/0 (site C75.1 only), 9580/0, 9582/0 |
| Craniopharyngioma | 9350-9352 | *None* | 9350/1, 9351/1, 9352/1 |
| *Unclassified Tumors* | | | |
| Hemangioma | 9121-9123, 9133, 9140 | 9133/3, 9140/3 | 9121/0, 9122/0, 9123/0, 9133/1 |
| Neoplasm, unspecified | 8000-8005, 8010, 8020, 8021 | 8000/3, 8001/3, 8002/3, 8003/3, 8004/3, 8005/3, 8010/3, 8020/3, 8021/3 | 8000/0,1, 8001/0,1, 8005/0, 8010/0 |
| All other | 8320, 8452, 8713, 8896, 8980, 9084/0, 9173, 9363, 9503 | 8320/3, 8452/3, 8896/3, 8980/3, 9503/3 | 8452/1, 8713/0, 9084/0, 9173/0, 9363/0 |

[i] *International Classification of Diseases for Oncology, 3rd Edition, 2000. World Health Organization, Geneva, Switzerland.*
[ii] *See the CBTRUS website for additional information about the specific histopathology codes included in each group: http://www.cbtrus.org.*
[iii] *Added starting with diagnosis year 2018.*
[iv] *This histopathology is re-coded from behavior /1 to /3 and included in estimates for malignant brain and other central nervous system tumors by cancer surveillance organizations. Please see the following for more information: Ostrom QT, Kruchko C, Barnholtz-Sloan JS. Pilocytic astrocytomas: where do they belong in cancer reporting? Neuro Oncol. 2020;22(2):298-300. doi: 10.1093/neuonc/noz202.*
[v] *Includes tumors formerly classified as primitive neuroectodermal tumors of the central nervous system (PNET).*
** All or some of this histopathology is included in the CBTRUS definition of gliomas, including ICD-O-3 histopathology codes 9380-9384, 9391-9460.*
***Abbreviations:*** *WHO, World Health Organization; NOS, not otherwise specified.*

### Supplementary Table 2. Individual ICD-O-3.2[i] histology codes reclassified as part of the revision process

| CBTRUS Tumor Type (CNS4/CNS4R)[ii] | CBTRUS Tumor Type (CNS5) | ICD-O-3.2 code | ICD-O-3.2 Histology label | Included sites | Included BMM | Total case count (2018-2022) |
|---|---|---|---|---|---|---|
| Diffuse astrocytoma | Astrocytoma, IDHmut | 9400/3 | Astrocytoma, NOS | C70-C72, C75.1- C75.3 | 01 | 3,627 |
| | Astrocytoma, NOS | 9400/3 | Astrocytoma, NOS | C70-C72, C75.1- C75.3 | 02,86,87,88,99 | 3,959 |
| | | 9410/3 | Protoplasmic astrocytoma | C70-C72, C75.1- C75.3 | n/a | <16 |
| | | 9411/3 | Gemistocytic astrocytoma | C70-C72, C75.1- C75.3 | n/a | 192 |
| | | 9420/3 | Fibrillary astrocytoma | C70-C72, C75.1- C75.3 | n/a | 59 |
| | | 9442/1 | Gliofibroma | C70-C72, C75.1- C75.3 | n/a | <16 |
| | Glioma, NOS | 9381/3 | Gliomatosis cerebri | C70-C72, C75.1- C75.3 | n/a | 85 |
| Anaplastic astrocytoma | Astrocytoma, IDHmut | 9401/3 | Astrocytoma, anaplastic | C70-C72, C75.1- C75.3 | 03 | 2,572 |
| | Astrocytoma, NOS | 9401/3 | Astrocytoma, anaplastic | C70-C72, C75.1- C75.3 | 04,86,87,88,99 | 2,658 |
| Unique astrocytoma variants | Pediatric-type diffuse low-grade gliomas | 9431/1 | Angiocentric glioma | C70-C72, C75.1- C75.3 | n/a | 139 |
| | Pleomorphic xanthoastrocytoma | 9424/3 | Pleomorphic xanthoastrocytoma | C70-C72, C75.1- C75.3 | n/a | 513 |
| | Subependymal giant cell astrocytoma | 9384/1 | Supependymal giant cell astrocytoma | C70-C72, C75.1- C75.3 | n/a | 335 |
| Glioblastoma | Astrocytoma, IDHmut | 9445/3 | Glioblastoma, IDH-mutant | C70-C72, C75.1- C75.3 | n/a | 1,477 |
| | Glioblastoma, IDHwt | 9440/3 | Glioblastoma, NOS | C70-C72, C75.1- C75.3 | 05 | 52,492 |
| | Glioblastoma, NOS | 9440/3 | Glioblastoma, NOS | C70-C72, C75.1- C75.3 | n/a | 12,168 |
| | | 9441/3 | Giant cell glioblastoma | C70-C72, C75.1- C75.3 | n/a | 333 |
| | | 9442/3 | Gliosarcoma | C70-C72, C75.1- C75.3 | n/a | 1,184 |
| Oligodendroglioma | Oligodendroglioma, IDHmut and 1p/19q-codel | 9450/3 | Oligodendroglioma, NOS | C70-C72, C75.1- C75.3 | 06 | 3,433 |
| | Oligodendroglial tumors, NOS | 9450/3 | Oligodendroglioma, NOS | C70-C72, C75.1- C75.3 | n/a | 337 |
| Anaplastic oligodendroglioma | Oligodendroglioma, IDHmut and 1p/19q-codel | 9451/3 | Oligodendroglioma, anaplastic | C70-C72, C75.1- C75.3 | 07 | 1,400 |
| | Oligodendroglial tumors, NOS | 9451/3 | Oligodendroglioma, anaplastic | C70-C72, C75.1- C75.3 | n/a | 99 |
| | | 9460/3 | Oligodendroblastoma | C70-C72, C75.1- C75.3 | n/a | <16 |
| Oligoastrocytic tumors | | 9382/3 | Mixed glioma | C70-C72, C75.1- C75.3 | n/a | 86 |
| Ependymal tumors | Myxopapillary ependymoma | 9394/1 | Myxopapillary ependymoma | C70-C72, C75.1- C75.3 | 07 | 1,546 |
| | Subependymoma | 9383/1 | Subependymoma | C70-C72, C75.1- C75.3 | 07 | 1,803 |
| | Spinal ependymoma | 9391/1 | Sellar ependymoma | C70.1, C72.0, C72.1 | n/a | <16 |
| | | 9391/3 | Ependymoma, NOS | C70.1, C72.0, C72.1 | n/a | 1,579 |
| | | 9392/3 | Ependymoma, anaplastic | C70.1, C72.0, C72.1 | n/a | 47 |
| | | 9393/3 | Papillary ependymoma | C70.1, C72.0, C72.1 | n/a | 25 |
| | | 9396/3 | Ependymoma, RELA fusion-positive | C70.1, C72.0, C72.1 | n/a | <16 |
| | Supratentorial ependymoma | 9391/1 | Sellar ependymoma | C71.0-C71.5 | n/a | <16 |
| | | 9391/3 | Ependymoma, NOS | C71.0-C71.5 | n/a | 421 |
| | | 9392/3 | Ependymoma, anaplastic | C71.0-C71.5 | n/a | 235 |
| | | 9393/3 | Papillary ependymoma | C71.0-C71.5 | n/a | <16 |

| | | | | | | |
|---|---|---|---|---|---|---|
| | | 9396/3 | Ependymoma, RELA fusion-positive | C71.0-C71.5 | n/a | 55 |
| | Posterior fossa ependymoma | 9391/1 | Sellar ependymoma | C71.6, C71.7 | n/a | <16 |
| | | 9391/3 | Ependymoma, NOS | C71.6, C71.7 | n/a | 580 |
| | | 9392/3 | Ependymoma, anaplastic | C71.6, C71.7 | n/a | 171 |
| | | 9393/3 | Papillary ependymoma | C71.6, C71.7 | n/a | <16 |
| | | 9396/3 | Ependymoma, RELA fusion-positive | C71.6, C71.7 | n/a | 23 |
| | Ependymoma, NOS | 9391/1 | Sellar ependymoma | C70.0, C70.9, C71.8, C71.9, C72.8, C72.9, C75.1- C75.3 | n/a | <16 |
| | | 9391/3 | Ependymoma, NOS | C70.0, C70.9, C71.8, C71.9, C72.8, C72.9, C75.1- C75.3 | n/a | 317 |
| | | 9392/3 | Ependymoma, anaplastic | C70.0, C70.9, C71.8, C71.9, C72.8, C72.9, C75.1- C75.3 | n/a | 130 |
| | | 9393/3 | Papillary ependymoma | C70.0, C70.9, C71.8, C71.9, C72.8, C72.9, C75.1- C75.3 | n/a | <16 |
| | | 9396/3 | Ependymoma, RELA fusion-positive | C70.0, C70.9, C71.8, C71.9, C72.8, C72.9, C75.1- C75.3 | n/a | 23 |
| Glioma malignant, NOS | Pediatric-type diffuse high-grade gliomas | 9385/3 | Diffuse midline glioma, H3 K27M-mutant | C70-C72, C75.1- C75.3 | n/a | 1,303 |
| | Glioma, NOS | 9380/3 | Glioma, malignant | C70-C72, C75.1- C75.3 | n/a | 8,415 |
| Other neuroepithelial tumors | Other circumscribed astrocytic gliomas | 9430/3 | Astroblastoma | C70-C72, C75.1- C75.3 | n/a | 58 |
| | | 9444/1 | Chordoid glioma | C70-C72, C75.1- C75.3 | n/a | 53 |
| | Glioma, NOS | 9423/3 | Polar spongioblastoma | C70-C72, C75.1- C75.3 | n/a | <16 |
| Neuronal and mixed neuronal-glial tumors | Ganglioglioma | 9505/1 | Ganglioglioma, NOS | C70-C72, C75.1- C75.3 | n/a | 1,910 |
| | | 9505/3 | Ganglioglioma, anaplastic | C70-C72, C75.1- C75.3 | n/a | 109 |
| | Desmoplastic infantile ganglioglioma/desmoplastic infantile astrocytoma | 9412/1 | Desmoplastic infantile astrocytoma | C70-C72, C75.1- C75.3 | n/a | 80 |
| | Dysembryoplastic neuroepithelial tumor | 9413/0 | Dysembryoplastic neuroepithelial tumor | C70-C72, C75.1- C75.3 | n/a | 1,149 |
| | Glioneuronal tumors | 9509/0 | Multinodular and vacuolating neuronal tumor | C71.1-Frontal lobe | Benign or borderline tumor | <16 |
| | | 9509/1 | Papillary glioneuronal tumor | C70-C72, C75.1- C75.3 | n/a | 374 |
| | | 9509/3 | Diffuse leptomeningeal glioneuronal tumor | C70-C72, C75.1- C75.3 | n/a | <16 |
| | Gangliocytoma | 9492/0 | Gangliocytoma | C70-C72, C75.1- C75.3 | n/a | 58 |
| | | 9493/0 | Dysplastic gangliocytoma of cerebellum (Lhermitte-Duclos) | C70-C72, C75.1- C75.3 | n/a | 72 |
| | Neurocytoma | 9506/1 | Centrol neurocytoma | C70-C72, C75.1- C75.3 | n/a | 511 |
| | Neuroblastoma | 9522/3 | Olfactory neuroblastoma | C30.0-Nasal cavity | n/a | 751 |
| | | 9523/3 | | C30.0-Nasal cavity | n/a | <16 |
| | Other neuronal and mixed neuronal-glial tumors | 8690/3 | Middle ear paraganglioma | C70-C72, C75.1- C75.3 | n/a | <16 |
| | | 9490/0 | Ganglioneuroma | C70-C72, C75.2- C75.3 | n/a | 84 |

| | | | | | | |
|---|---|---|---|---|---|---|
| | | 9490/3 | Ganglioneuroblastoma | C70-C72, C75.1- C75.3 | n/a | 30 |
| | Cauda equina neuroendocrine tumor | 8680/3 | Paraganglioma, malignant | C70-C72, C75.1- C75.3 | n/a | 38 |
| | | 8681/3 | Sympathetic paraganglioma | C70-C72, C75.1- C75.3 | n/a | <16 |
| | | 8693/3 | Extra-adrenal paraganglioma, malignant | C70-C72, C75.1- C75.3 | n/a | 53 |
| Tumors of the pineal region | Pineal tumors | 9360/1 | Pinealoma, NOS | C70-C72, C75.1- C75.3 | n/a | 41 |
| | | 9361/1 | Pineocytoma | C70-C72, C75.1- C75.3 | n/a | 227 |
| | | 9362/3 | Pineoblastoma | C70-C72, C75.1- C75.3 | n/a | 389 |
| | | 9395/3 | Papillary tumor of pineal region | C70-C72, C75.1- C75.3 | n/a | 69 |
| Embryonal tumors | Medulloblastoma, SHH-activated and TP53mut | 9476/3 | Medulloblastoma, SHH-activated and TP53-mutant | C70-C72, C75.1- C75.3 | n/a | 63 |
| | Medulloblastoma, non-WNT/non-SHH | 9477/3 | Medulloblastoma, non-WNT/non-SHH | C70-C72, C75.1- C75.3 | n/a | 404 |
| | Medulloblastoma, WNT-activated | 9475/3 | Medulloblastoma, WNT-activated | C70-C72, C75.1- C75.3 | n/a | 121 |
| | Medulloblastoma, SHH-activated and TP53wt | 9471/3 | Desmoplastic medulloblastoma | C70-C72, C75.1- C75.3 | 08 | 469 |
| | Medulloblastoma, NOS | 9470/3 | Medulloblastoma, NOS | C70-C72, C75.1- C75.3 | n/a | 937 |
| | | 9471/3 | Desmoplastic medulloblastoma | C70-C72, C75.1- C75.3 | 86,87,88,99 | 45 |
| | | 9472/3 | Medullomyoblastoma | C70-C72, C75.1- C75.3 | n/a | <16 |
| | | 9474/3 | Large cell medulloblastoma | C70-C72, C75.1- C75.3 | n/a | 130 |
| | Atypical teratoid/rhabdoid tumor | 9508/3 | Atypical teratoid/rhabdoid tumor | C70-C72, C75.1- C75.3 | n/a | 398 |
| | Embryonal tumor with multilayered rosettes | 9478/3 | Embryonal tumor with multilayered rosettes, NOS | C70-C72, C75.1- C75.3 | 09 | 97 |
| | Other CNS embryonal tumors | 8963/3 | Malignant rhabdoid tumor | C70-C72, C75.1- C75.3 | n/a | <16 |
| | | 9473/3 | Primitive neuroectodermal tumor | C70-C72, C75.1- C75.3 | n/a | 135 |
| | | 9500/3 | Neuroblastoma, NOS | C70-C72, C75.1- C75.3 | n/a | 174 |
| | | 9501/3 | Medulloepithelioma, NOS | C70-C72, C75.1- C75.3 | n/a | <16 |
| | Soft tissue tumor of uncertain differentiation | 9364/3 | Peripheral neuroectodermal tumor | C70-C72, C75.1- C75.3 | n/a | 29 |
| | | 9480/3 | Cerebellar sarcoma, NOS | C70-C72, C75.1- C75.3 | n/a | <16 |
| Nerve sheath tumors | Schwannoma | 9560/0 | Neurilemoma, NOS | C70-C72, C75.1- C75.3 | n/a | 38,118 |
| | | 9560/1 | Melanotic schwannoma | C70-C72, C75.1- C75.3 | n/a | 357 |
| | | 9560/3 | Neurilemmoma, malignant | C70-C72, C75.1- C75.3 | n/a | 107 |
| | Neurofibroma | 9540/0 | Neurofibroma, NOS | C70-C72, C75.1- C75.3 | n/a | 469 |
| | | 9541/0 | Melanotic neurofibroma | C70-C72, C75.1- C75.3 | n/a | <16 |
| | | 9550/0 | Plexiform neurofibroma | C70-C72, C75.1- C75.3 | n/a | 55 |
| | Malignant peripheral nerve sheath tumor | 9540/3 | Malignant peripheral nerve sheath tumor | C70-C72, C75.1- C75.3 | n/a | 116 |
| | | 9561/3 | MPNST with rhabdomyoblastic differentiation | C70-C72, C75.1- C75.3 | n/a | <16 |
| | Neuroma, NOS | 9570/0 | Neuroma, NOS | C70-C72, C75.1- C75.3 | n/a | 248 |
| | Other tumors of cranial and paraspinal nerves | 9571/0 | Perineurioma, NOS | C70-C72, C75.1- C75.3 | n/a | <16 |
| Other tumors of cranial and spinal nerves | Hybrid nerve sheath tumor | 9563/0 | Nerve sheath tumor, NOS | C70-C72, C75.1- C75.3 | n/a | 29 |
| | Other tumors of cranial and paraspinal nerves | 9562/0 | Neurothekeoma | C70-C72, C75.1- C75.3 | n/a | 24 |
| Meningiomas | Grade 1 meningioma | 9530/0 | Meningioma, NOS | C70-C72, C75.1- C75.3 | n/a | 129,882 |
| | | 9531/0 | Meningothelial meningioma | C70-C72, C75.1- C75.3 | n/a | 6,137 |

| | | | | | | |
|---|---|---|---|---|---|---|
| | | 9532/0 | Fibrous meningioma | C70-C72, C75.1- C75.3 | n/a | 1,519 |
| | | 9533/0 | Psammomatous meningioma | C70-C72, C75.1- C75.3 | n/a | 1,787 |
| | | 9534/0 | Angiomatous meningioma | C70-C72, C75.1- C75.3 | n/a | 809 |
| | | 9535/0 | Hemangioblastic meningioma | C70-C72, C75.1- C75.3 | n/a | <16 |
| | | 9537/0 | Transitional meningioma | C70-C72, C75.1- C75.3 | n/a | 2,261 |
| | Grade 2 meningioma (atypical) | 9530/0 | Meningioma, NOS | C70-C72, C75.1- C75.3 | n/a | 1,560 |
| | | 9531/0 | Meningothelial meningioma | C70-C72, C75.1- C75.3 | n/a | 123 |
| | | 9532/0 | Fibrous meningioma | C70-C72, C75.1- C75.3 | n/a | 21 |
| | | 9533/0 | Psammomatous meningioma | C70-C72, C75.1- C75.3 | n/a | <16 |
| | | 9534/0 | Angiomatous meningioma | C70-C72, C75.1- C75.3 | n/a | <16 |
| | | 9535/0 | Hemangioblastic meningioma | C70-C72, C75.1- C75.3 | n/a | <16 |
| | | 9537/0 | Transitional meningioma | C70-C72, C75.1- C75.3 | n/a | 58 |
| | | 9538/1 | Clear cell meningioma | C70-C72, C75.1- C75.3 | n/a | 661 |
| | | 9539/1 | Atypical meningioma | C70-C72, C75.1- C75.3 | n/a | 8,886 |
| | Grade 3 meningioma (malignant) | 9530/0 | Meningioma, NOS | C70-C72, C75.1- C75.3 | n/a | 71 |
| | | 9530/3 | Meningioma, malignant | C70-C72, C75.1- C75.3 | n/a | 1,573 |
| | | 9531/0 | Meningothelial meningioma | C70-C72, C75.1- C75.3 | n/a | <16 |
| | | 9533/0 | Psammomatous meningioma | C70-C72, C75.1- C75.3 | n/a | <16 |
| | | 9534/0 | Angiomatous meningioma | C70-C72, C75.1- C75.3 | n/a | <16 |
| | | 9535/0 | Hemangioblastic meningioma | C70-C72, C75.1- C75.3 | n/a | <16 |
| | | 9537/0 | Transitional meningioma | C70-C72, C75.1- C75.3 | n/a | <16 |
| | | 9538/3 | Papillary meningioma | C70-C72, C75.1- C75.3 | n/a | 76 |
| | | 9539/3 | Meningeal sarcomatosis | C70-C72, C75.1- C75.3 | n/a | 23 |
| | Ungraded meningioma | 9530/0 | Meningioma, NOS | C70-C72, C75.1- C75.3 | n/a | 52,097 |
| | | 9531/0 | Meningothelial meningioma | C70-C72, C75.1- C75.3 | n/a | 399 |
| | | 9532/0 | Fibrous meningioma | C70-C72, C75.1- C75.3 | n/a | 116 |
| | | 9533/0 | Psammomatous meningioma | C70-C72, C75.1- C75.3 | n/a | 141 |
| | | 9534/0 | Angiomatous meningioma | C70-C72, C75.1- C75.3 | n/a | 44 |
| | | 9537/0 | Transitional meningioma | C70-C72, C75.1- C75.3 | n/a | 162 |
| Mesenchymal tumors | Solitary fibrous tumor | 8815/0 | Solitary fibrous tumor | C70-C72, C75.1- C75.3 | n/a | 192 |
| | | 8815/1 | Solitary fibrous tumor/hemangiopericytoma Grade 2 | C70-C72, C75.1- C75.3 | n/a | 219 |
| | | 8815/3 | Solitary fibrous tumor, malignant | C70-C72, C75.1- C75.3 | n/a | 293 |
| | Hemangioma and vascular tumors | 9120/0 | Hemangioma, NOS | C70-C72, C75.1- C75.3 | n/a | 1,100 |
| | | 9120/3 | Hemangiosarcoma | C70-C72, C75.1- C75.3 | n/a | <16 |
| | | 9131/0 | Capillary hemangioma | C70-C72, C75.1- C75.3 | n/a | 53 |
| | | 9133/3 | Epithelioid hemangioendothelioma, malignant | C70-C72, C75.1- C75.3 | n/a | <16 |
| | | 9161/0 | Acquired tufted hemangioma | C70-C72, C75.1- C75.3 | n/a | 18 |
| | Hemangioblastoma | 9161/1 | Hemangioblastoma | C70-C72, C75.1- C75.3 | n/a | 2,949 |
| | Rhabdomyosarcoma | 8900/3 | Rhabdomyosarcoma, NOS | C70-C72, C75.1- C75.3 | n/a | <16 |
| | | 8901/3 | Pleomorphic rhabdomyosarcoma, adult type | C70-C72, C75.1- C75.3 | n/a | <16 |
| | | 8910/3 | Embryonal rhabdomyosarcoma | C70-C72, C75.1- C75.3 | n/a | <16 |
| | | 8912/3 | Spindle cell rhabdomyosarcoma | C70-C72, C75.1- C75.3 | n/a | <16 |

| | | | | | | |
|---|---|---|---|---|---|---|
| | | 8920/3 | Alveolar rhabdomyosarcoma | C70-C72, C75.1- C75.3 | n/a | <16 |
| | | 8921/3 | Rhabdomyosarcoma with ganglionic differentiation | C70-C72, C75.1- C75.3 | n/a | <16 |
| | Notochordal tumors | 9370/0 | Benign notochordal cell tumor | C70-C72, C75.1- C75.3 | n/a | <16 |
| | | 9370/3 | Chordoma, NOS | C70-C72, C75.1- C75.3 | n/a | 130 |
| | | 9371/3 | Chondroid chordoma | C70-C72, C75.1- C75.3 | n/a | <16 |
| | | 9372/3 | Dedifferentiated chordoma | C70-C72, C75.1- C75.3 | n/a | <16 |
| | Other mesenchymal tumors | 8711/0 | Glomus tumor, NOS | C70-C72, C75.1- C75.3 | n/a | <16 |
| | | 8711/1 | Glomangiomatosis | C70-C72, C75.1- C75.3 | n/a | <16 |
| | | 8711/3 | Glomus tumor, malignant | C70-C72, C75.1- C75.3 | n/a | <16 |
| | | 8800/0 | Soft tissue tumor, benign | C70-C72, C75.1- C75.3 | n/a | <16 |
| | | 8800/3 | Sarcoma, NOS | C70-C72, C75.1- C75.3 | n/a | 83 |
| | | 8801/3 | Spindle cell sarcoma | C70-C72, C75.1- C75.3 | n/a | 43 |
| | | 8802/3 | Giant cell sarcoma | C70-C72, C75.1- C75.3 | n/a | <16 |
| | | 8803/3 | Small cell sarcoma | C70-C72, C75.1- C75.3 | n/a | 19 |
| | | 8804/3 | Epithelioid sarcoma | C70-C72, C75.1- C75.3 | n/a | <16 |
| | | 8805/3 | Undifferentiated sarcoma | C70-C72, C75.1- C75.3 | n/a | <16 |
| | | 8806/3 | Desmoplastic small round cell tumor | C70-C72, C75.1- C75.3 | n/a | <16 |
| | | 8810/0 | Fibroma, NOS | C70-C72, C75.1- C75.3 | n/a | <16 |
| | | 8810/3 | Fibrosarcoma, NOS | C70-C72, C75.1- C75.3 | n/a | <16 |
| | | 8825/1 | Myofibroblastic tumor, NOS | C70-C72, C75.1- C75.3 | n/a | <16 |
| | | 8835/1 | Plexiform fibrohistiocytic tumor | C70-C72, C75.1- C75.3 | n/a | <16 |
| | | 8836/1 | Angiomatoid fibrous histiocytoma | C70-C72, C75.1- C75.3 | n/a | <16 |
| | | 8850/0 | Lipoma, NOS | C70-C72, C75.1- C75.3 | n/a | 320 |
| | | 8850/1 | Atypical lipoma | C70-C72, C75.1- C75.3 | n/a | <16 |
| | | 8851/0 | Fibrolipoma | C70-C72, C75.1- C75.3 | n/a | 96 |
| | | 8851/3 | Liposarcoma, well differentiated | C70-C72, C75.1- C75.3 | n/a | <16 |
| | | 8854/3 | Pleomorphic liposarcoma | C70-C72, C75.1- C75.3 | n/a | <16 |
| | | 8861/0 | Angiolipoma, NOS | C70-C72, C75.1- C75.3 | n/a | <16 |
| | | 8870/0 | Myelolipoma | C70-C72, C75.1- C75.3 | n/a | <16 |
| | | 8890/0 | Leiomyoma, NOS | C70-C72, C75.1- C75.3 | n/a | <16 |
| | | 8890/3 | Leiomyosarcoma, NOS | C70-C72, C75.1- C75.3 | n/a | <16 |
| | | 8897/1 | Smooth muscle tumor, NOS | C70-C72, C75.1- C75.3 | n/a | <16 |
| | | 8935/1 | Stromal tumor, NOS | C70-C72, C75.1- C75.3 | n/a | <16 |
| | | 8990/0 | Mesenchymoma, benign | C70-C72, C75.1- C75.3 | n/a | <16 |
| | | 8990/1 | Mesenchymoma, NOS | C70-C72, C75.1- C75.3 | n/a | <16 |
| | | 8990/3 | Mesenchymoma, malignant | C70-C72, C75.1- C75.3 | n/a | <16 |
| | | 9125/0 | Epithelioid hemangioma | C70-C72, C75.1- C75.3 | n/a | <16 |
| | | 9130/0 | Hemangioendothelioma, benign | C70-C72, C75.1- C75.3 | n/a | <16 |
| | | 9130/1 | Hemangioendothelioma, NOS | C70-C72, C75.1- C75.3 | n/a | <16 |
| | | 9150/0 | Hemangiopericytoma, benign | C70-C72, C75.1- C75.3 | n/a | <16 |
| | | 9150/1 | Hemangiopericytoma, NOS | C70-C72, C75.1- C75.3 | n/a | 49 |
| | | 9150/3 | Hemangiopericytoma, malignant | C70-C72, C75.1- C75.3 | n/a | 54 |

| | | | | | | |
|---|---|---|---|---|---|---|
| | | 9170/0 | Lymphangioma, NOS | C70-C72, C75.1- C75.3 | n/a | <16 |
| | | 9170/3 | Lymphangiosarcoma | C70-C72, C75.1- C75.3 | n/a | <16 |
| | | 9180/0 | Osteoma, NOS | C70-C72, C75.1- C75.3 | n/a | <16 |
| | | 9220/0 | Chondroma, NOS | C70-C72, C75.1- C75.3 | n/a | <16 |
| | | 9220/3 | Chondrosarcoma, NOS | C70-C72, C75.1- C75.3 | n/a | <16 |
| | | 9231/3 | Myxoid chondrosarcoma | C70-C72, C75.1- C75.3 | n/a | <16 |
| | | 9240/3 | Mesenchymal chondrosarcoma | C70-C72, C75.1- C75.3 | n/a | <16 |
| | | 9241/0 | Chondromyxoid fibroma | C70-C72, C75.1- C75.3 | n/a | <16 |
| | Non-langerhans cell histiocytosis | 8831/0 | Histiocytoma, NOS | C70-C72, C75.1- C75.3 | n/a | <16 |
| Primary melanocytic lesions | Melanocytic tumors | 8720/0 | Pigmented nevus, NOS | C70-C72, C75.1- C75.3 | n/a | <16 |
| | | 8720/3 | Malignant melanoma, NOS | C70-C72, C75.1- C75.3 | n/a | 90 |
| | | 8728/0 | Diffuse melanocytosis | C70-C72, C75.1- C75.3 | n/a | <16 |
| | | 8728/1 | Meningeal melanocytoma | C70-C72, C75.1- C75.3 | n/a | 48 |
| | | 8728/3 | Meningeal melanomatosis | C70-C72, C75.1- C75.3 | n/a | <16 |
| Lymphoma | Primary diffuse large B-cell lymphoma of the CNS | 9680/3 | ML, large B-cell, diffuse | C70-C72, C75.1- C75.3 | n/a | 6,905 |
| | Miscellaneous rare lymphomas in the CNS | 9699/3 | Marginal zone B-cell lymphoma, NOS | C70-C72, C75.1- C75.3 | n/a | 174 |
| | | 9702/3 | Mature T-cell lymphoma, NOS | C70-C72, C75.1- C75.3 | n/a | 65 |
| | | 9714/3 | Anaplastic large cell lymphoma, T-cell and Null cell type | C70-C72, C75.1- C75.3 | n/a | 21 |
| | | 9715/3 | Anaplastic large cell lymphoma, ALK negative | C70-C72, C75.1- C75.3 | n/a | <16 |
| | | 9719/3 | NK/T-cell lymphoma, nasal and nasal-type | C70-C72, C75.1- C75.3 | n/a | <16 |
| | Other CNS lymphoma | 9712/3 | Intravascular large B-cell lymphoma | C70-C72, C75.1- C75.3 | n/a | 31 |
| | Other malignant CNS lymphoma | 9590/3 | Malignant lymphoma, NOS | C70-C72, C75.1- C75.3 | n/a | 430 |
| | | 9591/3 | Malignant lymphoma, non-Hodgkin | C70-C72, C75.1- C75.3 | n/a | 471 |
| | Lymphoma, NOS | 9596/3 | Composite Hodgkin and non-Hodgkin lymphoma | C70-C72, C75.1- C75.3 | n/a | 23 |
| | | 9650/3 | Hodgkin lymphoma, NOS | C70-C72, C75.1- C75.3 | n/a | <16 |
| | | 9651/3 | Hodgkin lymphoma, lymphocyte-rich | C70-C72, C75.1- C75.3 | n/a | <16 |
| | | 9663/3 | Hodgkin lymphoma, nodular sclerosis, NOS | C70-C72, C75.1- C75.3 | n/a | <16 |
| | | 9671/3 | ML, lymphoplasmacytic | C70-C72, C75.1- C75.3 | n/a | 27 |
| | | 9673/3 | Mantle cell lymphoma | C70-C72, C75.1- C75.3 | n/a | <16 |
| | | 9680/1 | EBV positive mucocutaneous ulcer | C70-C72, C75.1- C75.3 | n/a | <16 |
| | | 9687/3 | Burkitt lymphoma, NOS | C70-C72, C75.1- C75.3 | n/a | 28 |
| | | 9688/3 | T-cell histiocyte rich large B-cell lymphoma | C70-C72, C75.1- C75.3 | n/a | <16 |
| | | 9690/3 | Follicular lymphoma, NOS | C70-C72, C75.1- C75.3 | n/a | 26 |
| | | 9691/3 | Follicular lymphoma, grade 2 | C70-C72, C75.1- C75.3 | n/a | 32 |
| | | 9695/1 | In situ follicular neoplasia | C70-C72, C75.1- C75.3 | n/a | <16 |
| | | 9695/3 | Follicular lymphoma, grade 1 | C70-C72, C75.1- C75.3 | n/a | <16 |
| | | 9698/3 | Follicular lymphoma, grade 3 | C70-C72, C75.1- C75.3 | n/a | <16 |
| | | 9735/3 | Plasmablastic lymphoma | C70-C72, C75.1- C75.3 | n/a | <16 |
| | | 9738/3 | Lrg B-cell lymphoma in HHV8-assoc. multicentric Castleman DZ | C70-C72, C75.1- C75.3 | n/a | <16 |

| | | | | | | |
|---|---|---|---|---|---|---|
| | | 9811/3 | B lymphoblastic leukemia/lymphoma, NOS | C70-C72, C75.1- C75.3 | n/a | 19 |
| | | 9818/3 | Leukemia/lymphoma with t(1;19)(q23;p13.3); E2A PBX1 (TCF3 PBX1) | C70-C72, C75.1- C75.3 | n/a | <16 |
| | | 9823/3 | Chronic lymphocytic leukemia/small lymphocytic lymphoma | C70-C72, C75.1- C75.3 | n/a | 25 |
| | | 9827/3 | Adult T-cell leukemia/lymphoma (HTLV-1 pos.) | C70-C72, C75.1- C75.3 | n/a | <16 |
| | | 9837/3 | T lymphoblastic leukemia/lymphoma | C70-C72, C75.1- C75.3 | n/a | <16 |
| | | 9930/3 | Myeloid sarcoma | C70-C72, C75.1- C75.3 | n/a | 29 |
| | | 9970/1 | Lymphoproliferative disorder, NOS | C70-C72, C75.1- C75.3 | n/a | <16 |
| | | 9971/1 | Post-transplant lymphoproliferative disorder, NOS | C70-C72, C75.1- C75.3 | n/a | 17 |
| | Langerhans cell histiocytosis | 9751/1 | Langerhans cell histiocytosis, NOS | C70-C72, C75.1- C75.3 | n/a | 35 |
| | | 9751/3 | Langerhans cell histiocytosis, NOS | C70-C72, C75.1- C75.3 | n/a | 66 |
| | Non-langerhans cell histiocytosis | 9755/3 | Histiocytic sarcoma | C70-C72, C75.1- C75.3 | n/a | 21 |
| | | 9756/3 | Langerhans cell sarcoma | C70-C72, C75.1- C75.3 | n/a | <16 |
| Other hematopoietic neoplasms | Other CNS lymphoma | 9766/1 | Angiocentric immunoproliferative lesion | C70-C72, C75.1- C75.3 | n/a | <16 |
| | | 9766/3 | Lymphomatoid granulomatosis, grade 3 | C70-C72, C75.1- C75.3 | n/a | <16 |
| | Non-langerhans cell histiocytosis | 9749/3 | Erdhiem-Chester Disease | C70-C72, C75.1- C75.3 | n/a | <16 |
| Germ cell tumors | Germinoma | 9064/3 | Germinoma | C70-C72, C75.1- C75.3 | n/a | 807 |
| | Non-germinoma | 8440/0 | Cystadenoma, NOS | C70-C72, C75.1- C75.3 | n/a | <16 |
| | | 9060/3 | Dysgerminoma | C70-C72, C75.1- C75.3 | n/a | <16 |
| | | 9061/3 | Seminoma, NOS | C70-C72, C75.1- C75.3 | n/a | <16 |
| | | 9065/3 | Germ cell tumor, nonseminomatous | C70-C72, C75.1- C75.3 | n/a | 25 |
| | | 9070/3 | Embryonal carcinoma, NOS | C70-C72, C75.1- C75.3 | n/a | 21 |
| | | 9071/3 | Yolk sac tumor | C70-C72, C75.1- C75.3 | n/a | <16 |
| | | 9080/0 | Teratoma, benign | C70-C72, C75.1- C75.3 | n/a | 103 |
| | | 9080/1 | Teratoma, NOS | C70-C72, C75.1- C75.3 | n/a | 36 |
| | | 9080/3 | Teratoma, malignant, NOS | C70-C72, C75.1- C75.3 | n/a | 41 |
| | | 9082/3 | Malignant teratoma, undiff. | C70-C72, C75.1- C75.3 | n/a | <16 |
| | | 9084/3 | Teratoma with malig. transformation | C70-C72, C75.1- C75.3 | n/a | <16 |
| | | 9085/3 | Mixed germ cell tumor | C70-C72, C75.1- C75.3 | n/a | 131 |
| | | 9100/3 | Choriocarcinoma | C70-C72, C75.1- C75.3 | n/a | <16 |
| | | 9101/3 | Choriocarcinoma combined w/ other germ cell elements | C70-C72, C75.1- C75.3 | n/a | <16 |
| Tumors of the pituitary | Gangliocytoma | 9492/0 | Gangliocytoma | C70-C72, C75.1- C75.3 | n/a | <16 |
| Hemangioma | Hemangioma and vascular tumors | 9121/0 | Cavernous hemangioma | C70-C72, C75.1- C75.3 | n/a | 4,571 |
| | | 9122/0 | Venous hemangioma | C70-C72, C75.1- C75.3 | n/a | 45 |
| | | 9123/0 | Racemose hemangioma | C70-C72, C75.1- C75.3 | n/a | <16 |
| All other | Melanocytic tumors | 9363/0 | Melanotic neuroectodermal tumor | C70-C72, C75.1- C75.3 | n/a | <16 |
| | Non-germinoma | 9084/0 | Dermoid cyst, NOS | C70-C72, C75.1- C75.3 | n/a | 301 |
| | Tumors of the pituitary gland | 8320/3 | Granular cell carcinoma | C70-C72, C75.1- C75.3 | n/a | <16 |
| | Choroid plexus tumors | 9390/0 | Choroid plexus papilloma, NOS | C70-C72, C75.1- C75.3 | n/a | 549 |

| | | | | | | |
|---|---|---|---|---|---|---|
| | | 9390/1 | Atypical choroid plexus papilloma | C70-C72, C75.1- C75.3 | n/a | 144 |
| | | 9390/3 | Choroid plexus papilloma, malignant | C70-C72, C75.1- C75.3 | n/a | 129 |

[i] *International Classification of Diseases for Oncology, 3rd Edition, 2000. World Health Organization, Geneva, Switzerland.*
[ii] *2021 WHO Classification of CNS tumors*
*-- Counts fewer than 16 were suppressed in accordance with data use policies.*
*Abbreviations: CBTRUS, Central Brain Tumor Registry of the United States; NOS, not otherwise specified.*
***Database**: CBTRUS US Cancer Statistics – CDC's National Program of Cancer Registries and NCI's Surveillance, Epidemiology, and End Results Program, 2018-2022.*

### Supplementary Table 3. Definition of specific tumor types by ICD-O-3.2 histology, behavior, topography, and brain molecular markers codes

| Tumor type | ICD-O-3 morphology and behavior code | Title of code and synonyms (from ICD-O-3.2 codebook) | Included ICD-O-3 topography codes | Brain molecular marker groups included (if applicable) | CNS WHO grade (if appliable) | Case Counts (2018-2022) |
|---|---|---|---|---|---|---|
| **Adult-type diffuse gliomas** | | | | | | |
| Astrocytoma, IDHmut | 9400/3 | Astrocytoma, NOS<br>*Synonyms: Astrocytic glioma; Astroglioma* | C70-C72, C75.1- C75.3 | 01 | n/a | 3,627 |
| | 9401/3 | Astrocytoma, anaplastic, NOS | C70-C72, C75.1- C75.3 | 03 | n/a | 2,572 |
| | 9445/3 | Glioblastoma, IDH-mutant<br>*Synonyms: Glioblastoma, secondary, IDH-mutant* | C70-C72, C75.1- C75.3 | n/a | n/a | 1,477 |
| Oligodendroglioma, IDHmut and 1p/19q-codel | 9450/3 | Oligodendroglioma, NOS | C70-C72, C75.1- C75.3 | 06 | n/a | 3,433 |
| | 9451/3 | Oligodendroglioma, anaplastic, NOS | C70-C72, C75.1- C75.3 | 07 | n/a | 1,400 |
| Astrocytoma, NOS | 9400/3 | Astrocytoma, NOS<br>*Synonyms: Astrocytic glioma; Astroglioma* | C70-C72, C75.1- C75.3 | 02,86,87,88,99 | n/a | 3,959 |
| | 9401/3 | Astrocytoma, anaplastic, NOS | C70-C72, C75.1- C75.3 | 04,86,87,88,99 | n/a | 2,658 |
| | 9410/3 | Protoplasmic astrocytoma | C70-C72, C75.1- C75.3 | n/a | n/a | <16 |
| | 9411/3 | Gemistocytic astrocytoma, NOS<br>*Synonyms: Gemistocytic astrocytoma, IDH-mutant; Gemistocytoma* | C70-C72, C75.1- C75.3 | n/a | n/a | 192 |
| | 9420/3 | Fibrillary astrocytoma<br>*Synonyms: Fibrous astrocytoma* | C70-C72, C75.1- C75.3 | n/a | n/a | 59 |
| | 9442/1 | Gliofibroma | C70-C72, C75.1- C75.3 | n/a | n/a | <16 |
| Glioblastoma, IDHwt | 9440/3 | Glioblastoma, NOS<br>*Synonyms: Glioblastoma multiforme; Spongioblastoma multiforme* | C70-C72, C75.1- C75.3 | 05 | n/a | 52,492 |
| Glioblastoma, NOS | 9440/3 | Glioblastoma, NOS<br>*Synonyms:* Glioblastoma multiforme; Spongioblastoma multiforme | C70-C72, C75.1- C75.3 | n/a | n/a | 12,168 |
| | 9441/3 | Giant cell glioblastoma<br>*Synonyms:* Monstrocellular sarcoma | C70-C72, C75.1- C75.3 | n/a | n/a | 333 |
| | 9442/3 | Gliosarcoma<br>*Synonyms:* Glioblastoma with sarcomatous component | C70-C72, C75.1- C75.3 | n/a | n/a | 1,184 |
| Oligodendroglial tumors, NOS | 9382/3 | Oligoastrocytoma, NOS<br>*Synonyms: Mixed glioma; Anaplastic oligoastrocytoma* | C70-C72, C75.1- C75.3 | n/a | n/a | 86 |
| | 9450/3 | Oligodendroglioma, NOS | C70-C72, C75.1- C75.3 | n/a | n/a | 337 |
| | 9451/3 | Oligodendroglioma, anaplastic, NOS | C70-C72, C75.1- C75.3 | n/a | n/a | 99 |
| | 9460/3 | Oligodendroblastoma | C70-C72, C75.1- C75.3 | n/a | n/a | <16 |
| **Pediatric-type diffuse low-grade gliomas** | 9431/1 | Angiocentric glioma | C70-C72, C75.1- C75.3 | n/a | n/a | 139 |
| **Pediatric-type diffuse high-grade gliomas** | 9385/3 | Diffuse midline glioma, H3 K27M-mutant | C70-C72, C75.1- C75.3 | n/a | n/a | 1,303 |

| | | | | | | |
|---|---|---|---|---|---|---|
| **Circumscribed astrocytic gliomas** | | | | n/a | | |
| Pilocytic astrocytoma | 9421/1 | Pilocytic astrocytoma<br>*Synonyms: Spongioblastoma, NOS; Juvenile astrocytoma; Piloid astrocytoma* | C70-C72, C75.1- C75.3 | n/a | n/a | 5,425 |
| | 9425/3 | Pilomyxoid astrocytoma | C70-C72, C75.1- C75.3 | n/a | n/a | 140 |
| Pleomorphic xanthoastrocytoma | 9424/3 | Pleomorphic xanthoastrocytoma, NOS | C70-C72, C75.1- C75.3 | n/a | n/a | 513 |
| Subependymal giant cell astrocytoma | 9384/1 | Subependymal giant cell astrocytoma | C70-C72, C75.1- C75.3 | n/a | n/a | 335 |
| Other circumscribed astrocytic gliomas | 9430/3 | Astroblastoma | C70-C72, C75.1- C75.3 | n/a | n/a | 58 |
| | 9444/1 | Chordoid glioma | C70-C72, C75.1- C75.3 | n/a | n/a | 53 |
| **Glioneuronal and neuronal tumors** | | | | n/a | | |
| Ganglioglioma | 9505/1 | Ganglioglioma, NOS<br>*Synonyms: Glioneuroma; Neuroastrocytoma* | C70-C72, C75.1- C75.3 | n/a | n/a | 1,910 |
| | 9505/3 | Ganglioglioma, anaplastic | C70-C72, C75.1- C75.3 | n/a | n/a | 109 |
| Desmoplastic infantile ganglioglioma/desmoplastic infantile astrocytoma | 9412/1 | Desmoplastic infantile astrocytoma | C70-C72, C75.1- C75.3 | n/a | n/a | 80 |
| Dysembryoplastic neuroepithelial tumor | 9413/0 | Dysembryoplastic neuroepithelial tumor | C70-C72, C75.1- C75.3 | n/a | n/a | 1,149 |
| Glioneuronal tumors | 9509/0 | Multinodular and vacuolating neuronal tumor | C70-C72, C75.1- C75.3 | n/a | | <16 |
| | 9509/1 | Papillary glioneuronal tumor | C70-C72, C75.1- C75.3 | n/a | n/a | 374 |
| | 9509/3 | Diffuse leptomeningeal glioneuronal tumor | C70-C72, C75.1- C75.3 | n/a | | <16 |
| Gangliocytoma | 9492/0 | Gangliocytoma, NOS | C70-C72, C75.1- C75.3 | n/a | n/a | 65 |
| | 9493/0 | Dysplastic gangliocytoma of cerebellum (Lhermitte-Duclos) | C70-C72, C75.1- C75.3 | n/a | n/a | 72 |
| Neurocytoma | 9506/1 | Central neurocytoma<br>*Synonyms: Neurocytoma, NOS; Lipomatous medulloblastoma; Medullocytoma; Neurolipocytoma* | C70-C72, C75.1- C75.3 | n/a | n/a | 511 |
| Neuroblastoma | 9522/3 | Olfactory neuroblastoma<br>*Synonyms: Esthesioneuroblastoma* | C30.0 | n/a | n/a | -- |
| | 9523/3 | Olfactory neuroepithelioma | C30.0 | n/a | n/a | <16 |
| Other neuronal and mixed neuronal-glial tumors | 8690/3 | Middle ear paraganglioma<br>*Synonyms: Glomus jugulare tumor, NOS; Jugular paraganglioma; Jugulotympanic paraganglioma* | C70-C72, C75.1- C75.3 | n/a | n/a | <16 |
| | 9490/0 | Ganglioneuroma | C70-C72, C75.1- C75.3 | n/a | n/a | 84 |
| | 9490/3 | Ganglioneuroblastoma | C70-C72, C75.1- C75.3 | n/a | n/a | 30 |
| **Ependymal tumors** | | | | n/a | | |
| Myxopapillary ependymoma | 9394/1 | Myxopapillary ependymoma | C70-C72, C75.1- C75.3 | n/a | n/a | 1,546 |
| Subependymoma | 9383/1 | Subependymoma<br>*Synonyms: Subependymal astrocytoma, NOS; Subependymal glioma* | C70-C72, C75.1- C75.3 | n/a | n/a | 1,803 |

| | | | | | | |
|---|---|---|---|---|---|---|
| Spinal ependymoma | 9391/3 | Ependymoma, NOS<br>*Synonyms: Epithelial ependymoma* | C70.1, C72.0, C72.1 | n/a | n/a | 1,579 |
| | 9392/3 | Ependymoma, anaplastic<br>*Synonyms: Ependymoblastoma* | C70.1, C72.0, C72.1 | n/a | n/a | 47 |
| | 9393/3 | Papillary ependymoma | C70.1, C72.0, C72.1 | n/a | n/a | 25 |
| | 9396/3 | Ependymoma, RELA fusion positive | C70.1, C72.0, C72.1 | n/a | n/a | <16 |
| Supratentorial ependymoma | 9391/3 | Ependymoma, NOS<br>*Synonyms: Epithelial ependymoma* | C71.0-C71.5 | n/a | n/a | 421 |
| | 9392/3 | Ependymoma, anaplastic<br>*Synonyms: Ependymoblastoma* | C71.0-C71.5 | n/a | n/a | 235 |
| | 9393/3 | Papillary ependymoma | C71.0-C71.5 | n/a | n/a | <16 |
| | 9396/3 | Ependymoma, RELA fusion positive | C71.0-C71.5 | n/a | n/a | 55 |
| Posterior fossa ependymoma | 9391/3 | Ependymoma, NOS<br>*Synonyms: Epithelial ependymoma* | C71.6, C71.7 | n/a | n/a | 580 |
| | 9392/3 | Ependymoma, anaplastic<br>*Synonyms: Ependymoblastoma* | C71.6, C71.7 | n/a | n/a | 171 |
| | 9393/3 | Papillary ependymoma | C71.6, C71.7 | n/a | n/a | <16 |
| | 9396/3 | Ependymoma, RELA fusion positive | C71.6, C71.7 | n/a | n/a | 23 |
| Ependymoma, NOS | 9391/3 | Ependymoma, NOS<br>*Synonyms: Epithelial ependymoma* | C70.0, C70.9, C71.8, C71.9, C72.8, C72.9, C75.1- C75.3 | n/a | n/a | 317 |
| | 9392/3 | Ependymoma, anaplastic<br>*Synonyms: Ependymoblastoma* | C70.0, C70.9, C71.8, C71.9, C72.8, C72.9, C75.1- C75.3 | n/a | n/a | 130 |
| | 9393/3 | Papillary ependymoma | C70.0, C70.9, C71.8, C71.9, C72.8, C72.9, C75.1- C75.3 | n/a | n/a | <16 |
| | 9396/3 | Ependymoma, RELA fusion positive | C70.0, C70.9, C71.8, C71.9, C72.8, C72.9, C75.1- C75.3 | n/a | n/a | 23 |
| **Glioma, NOS** | 9380/3 | Glioma, malignant<br>*Synonyms: Glioma, NOS* | C70-C72, C75.1- C75.3 | n/a | n/a | 8,415 |
| | 9381/3 | Gliomatosis cerebri | C70-C72, C75.1- C75.3 | n/a | n/a | 85 |
| | 9423/3 | Polar spongioblastoma<br>*Synonyms: Primitive polar spongioblastoma; Spongioblastoma polare* | C70-C72, C75.1- C75.3 | n/a | n/a | <16 |
| **Choroid plexus tumors** | 9390/0 | Choroid plexus papilloma, NOS | C71.5 | n/a | n/a | 549 |
| | 9390/1 | Atypical choroid plexus papilloma | C71.5 | n/a | n/a | 144 |
| | 9390/3 | Choroid plexus carcinoma<br>*Synonyms: Choroid plexus papilloma, anaplastic; Choroid plexus papilloma, malignant* | C71.5, C75.3 | n/a | n/a | 129 |
| **Embryonal tumors** | | | | n/a | | |
| Medulloblastoma, SHH-activated and TP53mut | 9476/3 | Medulloblastoma, SHH-activated and TP53-mutant | C70-C72, C75.1- C75.3 | n/a | n/a | 63 |

| | | | | | | |
|---|---|---|---|---|---|---|
| Medulloblastoma, non-WNT/non-SHH | 9477/3 | Medulloblastoma, non-WNT/non-SHH | C70-C72, C75.1- C75.3 | n/a | n/a | 404 |
| Medulloblastoma, WNT-activated | 9475/3 | Medulloblastoma, WNT-activated, NOS | C70-C72, C75.1- C75.3 | n/a | n/a | 121 |
| Medulloblastoma, SHH-activated and TP53wt | 9471/3 | Desmoplastic nodular medulloblastoma<br>*Synonyms: Circumscribed arachnoidal cerebellar sarcoma; Desmoplastic medulloblastoma, NOS* | C70-C72, C75.1- C75.3 | 08 | n/a | 469 |
| Medulloblastoma, NOS | 9470/3 | Medulloblastoma, NOS | C70-C72, C75.1- C75.3 | n/a | n/a | 937 |
| | 9471/3 | Desmoplastic nodular medulloblastoma<br>*Synonyms: Circumscribed arachnoidal cerebellar sarcoma; Desmoplastic medulloblastoma, NOS* | C70-C72, C75.1- C75.3 | 86,87,88,99 | n/a | 45 |
| | 9472/3 | Medullomyoblastoma | C70-C72, C75.1- C75.3 | n/a | n/a | <16 |
| | 9474/3 | Large cell medulloblastoma | C70-C72, C75.1- C75.3 | n/a | n/a | 130 |
| Atypical teratoid/rhabdoid tumor | 9508/3 | Atypical teratoid/rhabdoid tumor | C70-C72, C75.1- C75.3 | n/a | n/a | 398 |
| Embryonal tumor with multilayered rosettes | 9478/3 | Embryonal tumor with multilayered rosettes with C19MC alteration | C70-C72, C75.1- C75.3 | 09 | n/a | 97 |
| Other CNS embryonal tumors | 8963/3 | Rhabdoid tumor, NOS<br>*Synonyms:* Malignant rhabdoid tumor; Rhabdoid sarcoma | C70-C72, C75.1- C75.3 | n/a | n/a | <16 |
| | 9473/3 | CNS embryonal tumor, NOS<br>*Synonyms: Primitive neuroectodermal tumor, NOS; PNET, NOS; CPNET* | C70-C72, C75.1- C75.3 | n/a | n/a | 135 |
| | 9500/3 | Neuroblastoma, NOS<br>*Synonyms: Central neuroblastoma, CNS neuroblastoma, Sympathicoblastoma* | C70-C72, C75.1- C75.3 | n/a | n/a | 174 |
| | 9501/3 | Medulloepithelioma, NOS<br>*Synonyms: Diktyoma, malignant* | C70-C72, C75.1- C75.3 | n/a | n/a | <16 |
| **Pineal tumors** | 9360/1 | Pinealoma | C75.3 | n/a | n/a | 41 |
| | 9361/1 | Pineocytoma | C75.3 | n/a | n/a | 227 |
| | 9362/3 | Pineoblastoma<br>*Synonyms: Mixed pineocytoma-pineoblastoma* | C75.3 | n/a | n/a | 389 |
| | 9395/3 | Papillary tumor of pineal region | C71.9, C75.3, C71.5 | n/a | n/a | 69 |
| **Cranial and paraspinal nerve tumors** | | | | n/a | | |
| Schwannoma | 9560/0 | Schwannoma, NOS<br>*Synonyms: Neurilemoma, NOS; Neurinoma* | C70-C72, C75.1- C75.3 | n/a | n/a | 38,118 |
| | 9560/1 | Melanotic schwannoma<br>*Synonyms: Pigmented schwannoma* | C70-C72, C75.1- C75.3 | n/a | n/a | 357 |
| | 9560/3 | Neurilemoma, malignant<br>*Synonyms: Malignant schwannoma, NOS; Neurilemosarcoma* | C70-C72, C75.1- C75.3 | n/a | n/a | 107 |
| Neurofibroma | 9540/0 | Neurofibroma, NOS | C70-C72, C75.1- C75.3 | n/a | n/a | 469 |
| | 9541/0 | Melanotic neurofibroma | C70-C72, C75.1- C75.3 | n/a | n/a | <16 |
| | 9550/0 | Plexiform neurofibroma | C70-C72, C75.1- C75.3 | n/a | n/a | 55 |

| | | | | | | |
|---|---|---|---|---|---|---|
| | | *Synonyms:* Plexiform neuroma | | | | |
| Hybrid nerve sheath tumor | 9563/0 | Nerve sheath tumor, NOS | C70-C72, C75.1- C75.3 | n/a | n/a | 29 |
| Malignant peripheral nerve sheath tumor | 9540/3 | Malignant peripheral nerve sheath tumor, NOS<br>*Synonyms: MPNST, NOS; Neurofibrosarcoma; Neurogenic sarcoma; Neurosarcoma* | C70-C72, C75.1- C75.3 | n/a | n/a | 116 |
| | 9561/3 | Malignant peripheral nerve sheath tumor with rhabdomyoblastic differentiation<br>*Synonyms: Malignant schwannoma with rhabdomyoblastic differentiation; MPNST with rhabdomyoblastic differentiation; Triton tumor, malignant* | C70-C72, C75.1- C75.3 | n/a | n/a | <16 |
| Cauda equina neuroendocrine tumor | 8680/3 | Paraganglioma, NOS | C70-C72, C75.1- C75.3 | n/a | n/a | 38 |
| | 8681/3 | Sympathetic paraganglioma | C70-C72, C75.1- C75.3 | n/a | | <16 |
| | 8693/3 | Extra-adrenal paraganglioma, NOS | C70-C72, C75.1- C75.3 | n/a | n/a | 53 |
| Neuroma, NOS | 9570/0 | Neuroma, NOS | C70-C72, C75.1- C75.3 | n/a | n/a | 248 |
| Other tumors of cranial and paraspinal nerves | 9562/0 | Nerve sheath myxoma<br>*Synonyms: Neurothekeoma* | C70-C72, C75.1- C75.3 | n/a | n/a | 24 |
| | 9571/0 | Perineurioma, NOS | C70-C72, C75.1- C75.3 | n/a | n/a | <16 |
| **Meningiomas** | | | | n/a | | |
| Grade 1 meningioma | 9530/0 | Meningioma, NOS | C70-C72, C75.1- C75.3 | n/a | 1 | 129,882 |
| | 9531/0 | Meningothelial meningioma<br>*Synonyms: Endotheliomatous meningioma; Syncytial meningioma* | C70-C72, C75.1- C75.3 | n/a | 1 | 6,137 |
| | 9532/0 | Fibrous meningioma<br>*Synonyms: Fibroblastic meningioma* | C70-C72, C75.1- C75.3 | n/a | 1 | 1,519 |
| | 9533/0 | Psammomatous meningioma | C70-C72, C75.1- C75.3 | n/a | 1 | 1,787 |
| | 9534/0 | Angiomatous meningioma | C70-C72, C75.1- C75.3 | n/a | 1 | 809 |
| | 9535/0 | Hemangioblastic meningioma<br>*Synonyms: Angioblastic meningioma* | C70-C72, C75.1- C75.3 | n/a | 1 | <16 |
| | 9537/0 | Transitional meningioma<br>*Synonyms: Mixed meningioma* | C70-C72, C75.1- C75.3 | n/a | 1 | 2,261 |
| Grade 2 meningioma (atypical) | 9530/0 | Meningioma, NOS | C70-C72, C75.1- C75.3 | n/a | 2 | 1,560 |
| | 9531/0 | Meningothelial meningioma<br>*Synonyms: Endotheliomatous meningioma; Syncytial meningioma* | C70-C72, C75.1- C75.3 | n/a | 2 | 123 |
| | 9532/0 | Fibrous meningioma<br>*Synonyms: Fibroblastic meningioma* | C70-C72, C75.1- C75.3 | n/a | 2 | 21 |
| | 9533/0 | Psammomatous meningioma | C70-C72, C75.1- C75.3 | n/a | 2 | <16 |
| | 9534/0 | Angiomatous meningioma | C70-C72, C75.1- C75.3 | n/a | 2 | <16 |
| | 9535/0 | Hemangioblastic meningioma<br>*Synonyms: Angioblastic meningioma* | C70-C72, C75.1- C75.3 | n/a | 2 | <16 |
| | 9537/0 | Transitional meningioma<br>*Synonyms: Mixed meningioma* | C70-C72, C75.1- C75.3 | n/a | 2 | 58 |
| | 9538/1 | Clear cell meningioma | C70-C72, C75.1- C75.3 | n/a | Any | 661 |
| | 9539/1 | Atypical meningioma | C70-C72, C75.1- C75.3 | n/a | Any | 8,886 |

| | | | | | | |
|---|---|---|---|---|---|---|
| Grade 3 meningioma (malignant) | 9530/0 | Meningioma, NOS | C70-C72, C75.1- C75.3 | n/a | 3 or 4 | 71 |
| | 9530/3 | Meningioma, malignant<br>*Synonyms: Leptomeningeal sarcoma; Meningeal sarcoma; Meningioma, anaplastic; Meningothelial sarcoma* | C70-C72, C75.1- C75.3 | n/a | Any | 1,573 |
| | 9531/0 | Meningothelial meningioma<br>*Synonyms: Endotheliomatous meningioma; Syncytial meningioma* | C70-C72, C75.1- C75.3 | n/a | 3 or 4 | <16 |
| | 9533/0 | Psammomatous meningioma | C70-C72, C75.1- C75.3 | n/a | 3 or 4 | <16 |
| | 9534/0 | Angiomatous meningioma | C70-C72, C75.1- C75.3 | n/a | 3 or 4 | <16 |
| | 9535/0 | Hemangioblastic meningioma<br>*Synonyms: Angioblastic meningioma* | C70-C72, C75.1- C75.3 | n/a | 3 or 4 | <16 |
| | 9537/0 | Transitional meningioma<br>*Synonyms: Mixed meningioma* | C70-C72, C75.1- C75.3 | n/a | 3 or 4 | <16 |
| | 9538/3 | Papillary meningioma | C70-C72, C75.1- C75.3 | n/a | Any | 76 |
| | 9539/3 | Meningeal sarcomatosis | C70-C72, C75.1- C75.3 | n/a | Any | 23 |
| Ungraded meningioma | 9530/0 | Meningioma, NOS | C70-C72, C75.1- C75.3 | n/a | Missing | 52,097 |
| | 9531/0 | Meningothelial meningioma<br>*Synonyms: Endotheliomatous meningioma; Syncytial meningioma* | C70-C72, C75.1- C75.3 | n/a | Missing | 399 |
| | 9532/0 | Fibrous meningioma<br>*Synonyms: Fibroblastic meningioma* | | n/a | Missing | 116 |
| | 9533/0 | Psammomatous meningioma | C70-C72, C75.1- C75.3 | n/a | Missing | 141 |
| | 9534/0 | Angiomatous meningioma | C70-C72, C75.1- C75.3 | n/a | Missing | 44 |
| | 9537/0 | Transitional meningioma<br>*Synonyms: Mixed meningioma* | C70-C72, C75.1- C75.3 | n/a | Missing | 162 |
| **Mesenchymal, non-meningothelial tumors** | | | | n/a | | |
| Solitary fibrous tumor | 8815/0 | Solitary fibrous tumor/Hemangiopericytoma, grade 1<br>*Synonyms: Hemangiopericytoma, benign* | C70-C72, C75.1- C75.3 | n/a | n/a | 192 |
| | 8815/1 | Solitary fibrous tumor, NOS<br>*Synonyms: Hemangiopericytoma, NOS; Localized fibrous tumor* | C70-C72, C75.1- C75.3 | n/a | n/a | 219 |
| | 8815/3 | Solitary fibrous tumor, malignant<br>*Synonyms: Hemangiopericytoma, malignant* | C70-C72, C75.1- C75.3 | n/a | n/a | 293 |
| Hemangioma and vascular tumors | 9120/0 | Hemangioma, NOS<br>*Synonyms: Angioma, NOS; Chorioangioma* | C70-C72, C75.1- C75.3 | n/a | n/a | 1,100 |
| | 9120/3 | Hemangiosarcoma<br>*Synonyms: Angiosarcoma* | C70-C72, C75.1- C75.3 | n/a | n/a | <16 |
| | 9121/0 | Cavernous hemangioma | C70-C72, C75.1- C75.3 | n/a | n/a | 4,571 |
| | 9122/0 | Venous hemangioma | C70-C72, C75.1- C75.3 | n/a | n/a | 45 |
| | 9123/0 | Racemose hemangioma<br>*Synonyms: Arteriovenous hemangioma* | C70-C72, C75.1- C75.3 | n/a | n/a | <16 |
| | 9125/0 | Epithelioid hemangioma | | n/a | | <16 |

| | | | | | | |
|---|---|---|---|---|---|---|
| | | *Synonyms: Histiocytoid hemangioma* | | | | |
| | 9131/0 | Capillary hemangioma<br>*Synonyms: Hemangioma simplex; Infantile hemangioma; Juvenile hemangioma; Plexiform hemangioma* | C70-C72, C75.1- C75.3 | n/a | n/a | 53 |
| | 9133/3 | Epithelioid hemangioendothelioma, NOS<br>*Synonyms: Epithelioid hemangioendothelioma, malignant* | C70-C72, C75.1- C75.3 | n/a | n/a | <16 |
| | 9161/0 | Acquired tufted hemangioma<br>*Synonyms: Tufted hemangioma* | C70-C72, C75.1- C75.3 | n/a | n/a | 18 |
| Hemangioblastoma | 9161/1 | Hemangioblastoma<br>*Synonyms: Angioblastoma* | C70-C72, C75.1- C75.3 | n/a | n/a | 2,949 |
| Rhabdomyosarcoma | 8900/3 | Rhabdomyosarcoma, NOS<br>*Synonyms: Rhabdosarcoma* | C70-C72, C75.1- C75.3 | n/a | n/a | <16 |
| | 8901/3 | Pleomorphic rhabdomyosarcoma, adult type<br>*Synonyms: Rhabdomyosarcoma, pleomorphic type* | C70-C72, C75.1- C75.3 | n/a | n/a | <16 |
| | 8910/3 | Embryonal rhabdomyosarcoma, NOS<br>*Synonyms: Rhabdomyosarcoma, embryonal type; Botryoid sarcoma* | C70-C72, C75.1- C75.3 | n/a | n/a | <16 |
| | 8912/3 | Spindle cell rhabdomyosarcoma<br>*Synonyms: Sclerosing rhabdomyosarcoma; Rhabdomyosarcoma, spindle cell/sclerosing type* | C70-C72, C75.1- C75.3 | n/a | n/a | <16 |
| | 8920/3 | Alveolar rhabdomyosarcoma | C70-C72, C75.1- C75.3 | n/a | n/a | <16 |
| | 8921/3 | Ectomesenchymoma<br>*Synonyms: Rhabdomyosarcoma with ganglionic differentiation* | C70-C72, C75.1- C75.3 | n/a | n/a | <16 |
| Soft tissue tumor of uncertain differentiation | 9364/3 | Ewing sarcoma<br>*Synonyms: Ewing tumor; Peripheral neuroectodermal tumor; PPNET* | C70-C72, C75.1- C75.3 | n/a | | 29 |
| | 9480/3 | Cerebellar sarcoma, NOS | C70-C72, C75.1- C75.3 | n/a | | <16 |
| Notochordal tumors | 9370/0 | Benign notochordal cell tumor | C70-C72, C75.1- C75.3 | n/a | n/a | <16 |
| | 9370/3 | Chordoma, NOS | C70-C72, C75.1- C75.3 | n/a | n/a | 130 |
| | 9371/3 | Chondroid chordoma | C70-C72, C75.1- C75.3 | n/a | n/a | <16 |
| | 9372/3 | Dedifferentiated chordoma | C70-C72, C75.1- C75.3 | n/a | n/a | <16 |
| Other mesenchymal tumors | 8711/0 | Glomus tumor, NOS | C70-C72, C75.1- C75.3 | n/a | n/a | <16 |
| | 8711/1 | Glomangiomatosis | C70-C72, C75.1- C75.3 | n/a | n/a | <16 |
| | 8711/3 | Glomus tumor, malignant | C70-C72, C75.1- C75.3 | n/a | n/a | <16 |
| | 8800/0 | Soft tissue tumor, benign | C70-C72, C75.1- C75.3 | n/a | n/a | <16 |
| | 8800/3 | Sarcoma, NOS<br>*Synonyms:* Mesenchymal tumor, malignant; Soft tissue sarcoma; Soft tissue tumor, malignant | C70-C72, C75.1- C75.3 | n/a | n/a | 83 |
| | 8801/3 | Spindle cell sarcoma<br>*Synonyms:* Spindle cell sarcoma, undifferentiated | C70-C72, C75.1- C75.3 | n/a | n/a | 43 |
| | 8802/3 | Giant cell sarcoma<br>*Synonyms:* Pleomorphic cell sarcoma, NOS; Pleomorphic cell sarcoma, undifferentiated; Pleomorphic sarcoma | C70-C72, C75.1- C75.3 | n/a | n/a | <16 |

| | | | | | | |
|---|---|---|---|---|---|---|
| | 8803/3 | Small cell sarcoma<br>*Synonyms: Round cell sarcoma, NOS; Round cell sarcoma, undifferentiated* | C70-C72, C75.1- C75.3 | n/a | n/a | 19 |
| | 8804/3 | Epithelioid sarcoma, NOS<br>*Synonyms: Epithelioid cell sarcoma* | C70-C72, C75.1- C75.3 | n/a | n/a | <16 |
| | 8805/3 | Undifferentiated sarcoma | C70-C72, C75.1- C75.3 | n/a | n/a | <16 |
| | 8806/3 | Desmoplastic small round cell tumor | C70-C72, C75.1- C75.3 | n/a | n/a | <16 |
| | 8810/0 | Fibroma, NOS | C70-C72, C75.1- C75.3 | n/a | n/a | <16 |
| | 8810/3 | Fibrosarcoma, NOS | C70-C72, C75.1- C75.3 | n/a | n/a | <16 |
| | 8825/1 | Myofibroblastic tumor, NOS<br>*Synonyms: Inflammatory myofibroblastic tumor* | C70-C72, C75.1- C75.3 | n/a | n/a | <16 |
| | 8835/1 | Plexiform fibrohistiocytic tumor | C70-C72, C75.1- C75.3 | n/a | n/a | <16 |
| | 8836/1 | Angiomatoid fibrous histiocytoma | C70-C72, C75.1- C75.3 | n/a | n/a | <16 |
| | 8850/0 | Lipoma, NOS | C70-C72, C75.1- C75.3 | n/a | n/a | 320 |
| | 8850/1 | Atypical lipomatous tumor<br>*Synonyms: Atypical lipoma; Liposarcoma, well differentiated, superficial* | C70-C72, C75.1- C75.3 | n/a | n/a | <16 |
| | 8851/0 | Fibrolipoma | C70-C72, C75.1- C75.3 | n/a | n/a | 96 |
| | 8851/3 | Liposarcoma, well differentiated, NOS<br>*Synonyms: Lipoma-like liposarcoma; Liposarcoma, differentiated* | C70-C72, C75.1- C75.3 | n/a | n/a | <16 |
| | 8854/3 | Pleomorphic liposarcoma | C70-C72, C75.1- C75.3 | n/a | n/a | <16 |
| | 8861/0 | Angiolipoma, NOS | C70-C72, C75.1- C75.3 | n/a | n/a | <16 |
| | 8870/0 | Myelolipoma | C70-C72, C75.1- C75.3 | n/a | n/a | <16 |
| | 8890/0 | Leiomyoma, NOS<br>*Synonyms: Fibroid uterus; Fibromyoma; Leiomyofibroma; Lipomatous leiomyoma; Dissecting leiomyoma* | C70-C72, C75.1- C75.3 | n/a | n/a | <16 |
| | 8890/3 | Leiomyosarcoma, NOS | C70-C72, C75.1- C75.3 | n/a | n/a | <16 |
| | 8897/1 | Smooth muscle tumor of uncertain malignant potential<br>*Synonyms: Smooth muscle tumor, atypica* | C70-C72, C75.1- C75.3 | n/a | n/a | <16 |
| | 8935/1 | Stromal tumor, NOS | C70-C72, C75.1- C75.3 | n/a | n/a | <16 |
| | 8990/0 | Mesenchymoma, benign<br>*Synonyms: Phosphaturic mesenchymal tumor, NOS* | C70-C72, C75.1- C75.3 | n/a | n/a | <16 |
| | 8990/1 | Mesenchymoma, NOS<br>*Synonyms: Mixed mesenchymal tumor* | C70-C72, C75.1- C75.3 | n/a | n/a | <16 |
| | 8990/3 | Mesenchymoma, malignant<br>*Synonyms: Mixed mesenchymal sarcoma* | C70-C72, C75.1- C75.3 | n/a | n/a | <16 |
| | 9130/0 | Hemangioendothelioma, benign | C70-C72, C75.1- C75.3 | n/a | n/a | <16 |
| | 9130/1 | Hemangioendothelioma, NOS<br>*Synonyms: Angioendothelioma* | C70-C72, C75.1- C75.3 | n/a | n/a | <16 |
| | 9150/0 | Hemangiopericytoma, benign | | n/a | | <16 |
| | 9150/1 | Hemangiopericytoma, malignant | | n/a | | 49 |
| | 9150/3 | Hemangiopericytoma, NOS | | n/a | | 54 |

| | | | | | | |
|---|---|---|---|---|---|---|
| | 9170/0 | Lymphangioma, NOS<br>*Synonyms: Lymphangioendothelioma, NOS* | C70-C72, C75.1- C75.3 | n/a | n/a | <16 |
| | 9170/3 | Lymphangiosarcoma<br>*Synonyms: Lymphangioendothelial sarcoma; Lymphangioendothelioma, malignant* | C70-C72, C75.1- C75.3 | n/a | n/a | <16 |
| | 9180/0 | Osteoma, NOS | C70-C72, C75.1- C75.3 | n/a | n/a | <16 |
| | 9220/0 | Chondroma, NOS | C70-C72, C75.1- C75.3 | n/a | n/a | <16 |
| | 9220/3 | Chondrosarcoma, NOS | C70-C72, C75.1- C75.3 | n/a | n/a | <16 |
| | 9231/3 | Myxoid chondrosarcoma | C70-C72, C75.1- C75.3 | n/a | n/a | <16 |
| | 9240/3 | Mesenchymal chondrosarcoma | C70-C72, C75.1- C75.3 | n/a | n/a | <16 |
| | 9241/0 | Chondromyxoid fibroma | C70-C72, C75.1- C75.3 | n/a | n/a | <16 |
| Melanocytic tumors | 8720/0 | Pigmented nevus, NOS<br>*Synonyms: Nevus, NOS; Melanocytic nevus, NOS* | C70-C72, C75.1- C75.3 | n/a | n/a | <16 |
| | 8720/3 | Malignant melanoma, NOS<br>*Synonyms: Melanoma, NOS* | C70-C72, C75.1- C75.3 | n/a | n/a | 90 |
| | 8728/0 | Meningeal melanocytosis | C70-C72, C75.1- C75.3 | n/a | n/a | <16 |
| | 8728/1 | Meningeal melanocytoma | C70-C72, C75.1- C75.3 | n/a | n/a | 48 |
| | 8728/3 | Meningeal melanomatosis | C70-C72, C75.1- C75.3 | n/a | n/a | <16 |
| | 9363/0 | Melanotic neuroectodermal tumor<br>*Synonyms: Melanoameloblastoma; Melanotic progonoma; Retinal anlage tumor* | C70-C72, C75.1- C75.3 | n/a | n/a | <16 |
| **Hematolymphoid tumors** | | | | n/a | | |
| Primary diffuse large B-cell lymphoma of the CNS | 9680/3 | Diffuse large B-cell lymphoma, NOS<br>*Synonyms: Malignant lymphoma, large B-cell, diffuse, NOS; Malignant lymphoma, histiocytic, NOS; Malignant lymphoma, large B-cell, NOS; Malignant lymphoma, large B-cell, diffuse, centroblastic, NOS; Malignant lymphoma, large cell, NOS; Malignant lymphoma, large cell, cleaved, NOS; Malignant lymphoma, large cell, diffuse, NOS; Malignant lymphoma, large cell, noncleaved, NOS; Malignant lymphoma, noncleaved, NOS; Malignant lymphoma, noncleaved, diffuse, NOS; Malignant lymphoma, histiocytic, diffuse; Malignant lymphoma, large cell, cleaved and noncleaved; Malignant lymphoma, large cell, cleaved, diffuse; Malignant lymphoma, large cell, noncleaved, diffuse* | C70-C72, C75.1- C75.3 | n/a | n/a | 6,905 |
| Miscellaneous rare lymphomas in the CNS | 9699/3 | Marginal zone B-cell lymphoma, NOS<br>*Synonyms: Marginal zone lymphoma, NOS; BALT lymphoma; Bronchus-associated lymphoid tissue lymphoma; MALT lymphoma; Monocytoid B-cell lymphoma; Mucosa-associated lymphoid tissue lymphoma; Nodal marginal zone lymphoma; SALT lymphoma; Skin-associated lymphoid tissue lymphoma* | C70-C72, C75.1- C75.3 | n/a | n/a | 174 |
| | 9702/3 | Mature T-cell lymphoma, NOS | C70-C72, C75.1- C75.3 | n/a | n/a | 65 |

| | | | | | | |
|---|---|---|---|---|---|---|
| | | *Synonyms:* Peripheral T-cell lymphoma, NOS; T-cell lymphoma, NOS; Peripheral T-cell lymphoma, large cell; Peripheral T-cell lymphoma, pleomorphic medium and large cell; Peripheral T-cell lymphoma, pleomorphic small cell; T-zone lymphoma; Lennert lymphoma | | | | |
| | 9714/3 | Anaplastic large cell lymphoma, T-cell and Null-cell type *Synonyms:* Large cell (Ki-1 positive) lymphoma; Anaplastic large cell lymphoma, CD30 positive | C70-C72, C75.1- C75.3 | n/a | n/a | 21 |
| | 9715/3 | Anaplastic large cell lymphoma, ALK negative | C70-C72, C75.1- C75.3 | n/a | n/a | <16 |
| | 9719/3 | NK/T-cell lymphoma, nasal and nasal type *Synonyms:* Malignant reticulosis, NOS; Angiocentric T-cell lymphoma; Extranodal NK/T-cell lymphoma, nasal type; Malignant midline reticulosis; Polymorphic reticulosis; T/NK-cell lymphoma | C70-C72, C75.1- C75.3 | n/a | n/a | <16 |
| Other CNS lymphoma | 9712/3 | Intravascular large B-cell lymphoma Synonyms: Intravascular B-cell lymphoma; Angioendotheliomatosis; Angiotropic lymphoma | C70-C72, C75.1- C75.3 | n/a | n/a | 31 |
| | 9766/1 | Angiocentric immunoproliferative lesion | C70-C72, C75.1- C75.3 | n/a | n/a | <16 |
| | 9766/3 | Lymphomatoid granulomatosis, grade 3 | C70-C72, C75.1- C75.3 | n/a | n/a | <16 |
| Other malignant CNS lymphoma | 9590/3 | Malignant lymphoma, NOS *Synonyms:* Lymphoma, NOS; Microglioma | C70-C72, C75.1- C75.3 | n/a | n/a | 430 |
| | 9591/3 | Malignant lymphoma, non-Hodgkin, NOS *Synonyms:* Non-Hodgkin lymphoma, NOS; Lymphosarcoma, diffuse; Reticulosarcoma, NOS; Reticulum cell sarcoma, diffuse; Reticulosarcoma, diffuse; Malignant lymphoma, cleaved cell, NOS; Malignant lymphoma, small cleaved cell, NOS; Malignant lymphoma, undifferentiated cell type, NOS; Malignant lymphoma, undifferentiated cell, non-Burkitt | C70-C72, C75.1- C75.3 | n/a | n/a | 471 |
| Lymphoma, NOS | 9596/3 | Composite Hodgkin and non-Hodgkin lymphoma | C70-C72, C75.1- C75.3 | n/a | n/a | 23 |
| | 9650/3 | Hodgkin lymphoma, NOS *Synonyms:* Hodgkin disease, NOS; Malignant lymphoma, Hodgkin | C70-C72, C75.1- C75.3 | n/a | n/a | <16 |
| | 9651/3 | Hodgkin lymphoma, lymphocyte-rich *Synonyms:* Classical Hodgkin lymphoma, lymphocyte-rich; Hodgkin disease, lymphocyte predominance, diffuse; Hodgkin disease, lymphocytic-histiocytic predominance | C70-C72, C75.1- C75.3 | n/a | n/a | <16 |
| | 9663/3 | Hodgkin lymphoma, nodular sclerosis, NOS *Synonyms:* Classical Hodgkin lymphoma, nodular sclerosis, NOS; Hodgkin disease, nodular sclerosis, NOS | C70-C72, C75.1- C75.3 | n/a | n/a | <16 |
| | 9671/3 | Lymphoplasmacytic lymphoma *Synonyms:* Malignant lymphoma, lymphoplasmacytoid | C70-C72, C75.1- C75.3 | n/a | n/a | 27 |
| | 9673/3 | Mantle cell lymphoma | C70-C72, C75.1- C75.3 | n/a | n/a | <16 |

| | | | | | | |
|---|---|---|---|---|---|---|
| | | *Synonyms:* Malignant lymphoma, centrocytic; Malignant lymphoma, lymphocytic, intermediate differentiation, diffuse; Malignant lymphomatous polyposis; Mantle zone lymphoma | | | | |
| | 9680/1 | EBV positive mucocutaneous ulcer | C70-C72, C75.1- C75.3 | n/a | n/a | <16 |
| | 9687/3 | Burkitt lymphoma, NOS<br>*Synonyms:* Burkitt tumor; Malignant lymphoma, small noncleaved, Burkitt type; Malignant lymphoma, undifferentiated, Burkitt type; Burkitt cell leukemia; Acute leukemia, Burkitt type; Acute lymphoblastic leukemia, mature B-cell type; B-ALL; FAB L3 | C70-C72, C75.1- C75.3 | n/a | n/a | 28 |
| | 9688/3 | T-cell/histiocyte rich large B-cell lymphoma<br>*Synonyms:* T-cell rich large B-cell lymphoma; Histiocyte-rich large B-cell lymphoma | C70-C72, C75.1- C75.3 | n/a | n/a | <16 |
| | 9690/3 | Follicular lymphoma, NOS<br>*Synonyms:* Malignant lymphoma, follicle center, NOS; Malignant lymphoma, follicular, NOS; Malignant lymphoma, lymphocytic, nodular, NOS; Malignant lymphoma, nodular, NOS; Malignant lymphoma, centroblastic-centrocytic, follicular; Malignant lymphoma, follicle center, follicular | C70-C72, C75.1- C75.3 | n/a | n/a | 26 |
| | 9691/3 | Follicular lymphoma, grade 2<br>*Synonyms:* Malignant lymphoma, mixed cell type, follicular; Malignant lymphoma, mixed cell type, nodular; Malignant lymphoma, mixed lymphocytic-histiocytic, nodular; Malignant lymphoma, mixed small cleaved and large cell, follicular | C70-C72, C75.1- C75.3 | n/a | n/a | 32 |
| | 9695/1 | In situ follicular neoplasia<br>*Synonyms:* In situ follicular lymphoma | C70-C72, C75.1- C75.3 | n/a | n/a | <16 |
| | 9695/3 | Follicular lymphoma, grade 1<br>*Synonyms:* Follicular lymphoma, small cleaved cell; Malignant lymphoma, lymphocytic, poorly differentiated, nodular; Malignant lymphoma, small cleaved cell, follicular | C70-C72, C75.1- C75.3 | n/a | n/a | <16 |
| | 9698/3 | Follicular lymphoma, grade 3<br>*Synonyms:* Malignant lymphoma, large cell, follicular, NOS; Malignant lymphoma, noncleaved cell, follicular, NOS; Follicular lymphoma, grade 3A; Follicular lymphoma, grade 3B; Malignant lymphoma, centroblastic, follicular; Malignant lymphoma, histiocytic, nodular; Malignant lymphoma, large cell, noncleaved, follicular; Malignant lymphoma, large cleaved cell, follicular; Malignant lymphoma, lymphocytic, well differentiated, nodular | C70-C72, C75.1- C75.3 | n/a | n/a | <16 |
| | 9735/3 | Plasmablastic lymphoma | C70-C72, C75.1- C75.3 | n/a | n/a | <16 |

| | | | | | | |
|---|---|---|---|---|---|---|
| | 9738/3 | HHV8 positive diffuse large B-cell lymphoma *Synonyms:* Large B-cell lymphoma arising in HHV8-associated multicentric Castleman disease | C70-C72, C75.1- C75.3 | n/a | n/a | <16 |
| | 9811/3 | B lymphoblastic leukemia/lymphoma, NOS *Synonyms:* c-ALL; Common ALL; Common precursor B ALL; Pre-B ALL; Pre-pre-B ALL; Pro-B ALL; Precursor B-cell lymphoblastic lymphoma; Precursor B-cell lymphoblastic leukemia | C70-C72, C75.1- C75.3 | n/a | n/a | 19 |
| | 9818/3 | B lymphoblastic leukemia/lymphoma with t(1;19)(q23;p13.3); E2A-PBX1 (TCF3-PBX1) | C70-C72, C75.1- C75.3 | n/a | n/a | <16 |
| | 9823/3 | B-cell chronic lymphocytic leukemia/small lymphocytic lymphoma *Synonyms:* Chronic lymphatic leukemia; Chronic lymphocytic leukemia; Chronic lymphocytic leukemia, B-cell type; Chronic lymphoid leukemia; Malignant lymphoma, small B lymphocytic, NOS; Malignant lymphoma, lymphocytic, NOS; Malignant lymphoma, lymphocytic, diffuse, NOS; Malignant lymphoma, small cell, NOS; Malignant lymphoma, small lymphocytic, NOS; Malignant lymphoma, lymphocytic, well differentiated, diffuse; Malignant lymphoma, small cell diffuse; Malignant lymphoma, small lymphocytic, diffuse | C70-C72, C75.1- C75.3 | n/a | n/a | 25 |
| | 9827/3 | Adult T-cell leukemia/lymphoma (HTLV-1 positive) *Synonyms:* Adult T-cell leukemia; Adult T-cell lymphoma; Adult T-cell lymphoma/leukemia | C70-C72, C75.1- C75.3 | n/a | | <16 |
| | 9837/3 | Precursor T-cell lymphoblastic leukemia *Synonyms:* Cortical T ALL; Mature T ALL; Pre-T ALL; Pro-T ALL; Precursor T-cell lymphoblastic lymphoma | C70-C72, C75.1- C75.3 | n/a | | <16 |
| | 9930/3 | Myeloid sarcoma *Synonyms:* Chloroma; Granulocytic sarcoma | C70-C72, C75.1- C75.3 | n/a | n/a | 29 |
| | 9970/1 | Lymphoproliferative disorder, NOS *Synonyms:* Lymphoproliferative disease, NOS | C70-C72, C75.1- C75.3 | n/a | n/a | <16 |
| | 9971/1 | Post-transplant lymphoproliferative disorder, NOS *Synonyms:* PTLD, NOS | C70-C72, C75.1- C75.3 | n/a | n/a | 17 |
| Other hematopoietic neoplasms | 9734/3 | Plasmacytoma, extramedullary *Synonyms:* Extraosseous plasmacytoma | C70-C72, C75.1- C75.3 | n/a | n/a | 32 |
| | 9758/3 | Follicular dendritic cell sarcoma | C70-C72, C75.1- C75.3 | n/a | n/a | <16 |
| Langerhans cell histiocytosis | 9751/1 | Langerhans cell histiocytosis, NOS | C70-C72, C75.1- C75.3 | n/a | n/a | 35 |
| | 9751/3 | Langerhans cell histiocytosis, disseminated *Synonyms:* Langerhans cell granulomatosis | C70-C72, C75.1- C75.3 | n/a | n/a | 66 |
| Non-langerhans cell histiocytosis | 8831/0 | Histiocytoma, NOS *Synonyms: Deep benign fibrous histiocytoma; Reticulohistocytosis* | C70-C72, C75.1- C75.3 | n/a | | <16 |
| | 9749/3 | Erdheim-Chester disease | C70-C72, C75.1- C75.3 | n/a | n/a | <16 |

| | | | | | | |
|---|---|---|---|---|---|---|
| | 9755/3 | Histiocytic sarcoma | C70-C72, C75.1- C75.3 | n/a | n/a | 21 |
| | 9756/3 | Langerhans cell sarcoma | C70-C72, C75.1- C75.3 | n/a | n/a | <16 |
| **Germ cell tumors** | | | | n/a | | |
| Germinoma | 9064/3 | Germinoma<br>*Synonyms:* Germ cell tumor, NOS | C70-C72, C75.1- C75.3 | n/a | n/a | 807 |
| Non-germinoma | 8440/0 | Cystadenoma, NOS<br>*Synonyms:* Cystoma, NOS | C70-C72, C75.1- C75.3 | n/a | n/a | <16 |
| | 9060/3 | Dysgerminoma | C70-C72, C75.1- C75.3 | n/a | n/a | <16 |
| | 9061/3 | Seminoma, NOS | C70-C72, C75.1- C75.3 | n/a | n/a | <16 |
| | 9065/3 | Germ cell tumor, nonseminomatous | C70-C72, C75.1- C75.3 | n/a | n/a | 25 |
| | 9070/3 | Embryonal carcinoma, NOS<br>*Synonyms:* Embryonal adenocarcinoma | C70-C72, C75.1- C75.3 | n/a | n/a | 21 |
| | 9071/3 | Yolk sac tumor, NOS<br>*Synonyms:* Embryonal carcinoma, infantile; Yolk sac tumor, pre-pubertal type; Endodermal sinus tumor; Orchioblastoma; Polyvesicular vitelline tumor | C70-C72, C75.1- C75.3 | n/a | n/a | <16 |
| | 9080/0 | Teratoma, benign<br>*Synonyms:* Adult teratoma, NOS; Cystic teratoma, NOS; Adult cystic teratoma; Mature teratoma, NOS; Teratoma, differentiated | C70-C72, C75.1- C75.3 | n/a | n/a | 103 |
| | 9080/1 | Teratoma, NOS<br>*Synonyms:* Solid teratoma | C70-C72, C75.1- C75.3 | n/a | n/a | 36 |
| | 9080/3 | Teratoma, malignant, NOS<br>*Synonyms:* Embryonal teratoma; Teratoblastoma, malignant; Immature teratoma, NOS | C70-C72, C75.1- C75.3 | n/a | n/a | 41 |
| | 9082/3 | Malignant teratoma, undifferentiated<br>*Synonyms:* Malignant teratoma, anaplastic | C70-C72, C75.1- C75.3 | n/a | n/a | <16 |
| | 9084/0 | Dermoid cyst, NOS<br>*Synonyms:* Dermoid, NOS; Teratoma, mature, prepubertal type | C70-C72, C75.1- C75.3 | n/a | n/a | 301 |
| | 9084/3 | Teratoma with malignant transformation<br>*Synonyms:* Teratoma with somatic type malignancies; Dermoid cyst with malignant transformation; Dermoid cyst with secondary tumor | C70-C72, C75.1- C75.3 | n/a | n/a | <16 |
| | 9085/3 | Mixed germ cell tumor | C70-C72, C75.1- C75.3 | n/a | n/a | 131 |
| | 9100/3 | Choriocarcinoma, NOS<br>*Synonyms:* Chorioepithelioma; Chorionepithelioma | C70-C72, C75.1- C75.3 | n/a | n/a | <16 |
| | 9101/3 | Choriocarcinoma combined with other germ cell elements<br>*Synonyms:* Choriocarcinoma combined with embryonal carcinoma; Choriocarcinoma combined with teratoma | C70-C72, C75.1- C75.3 | n/a | n/a | <16 |
| **Tumors of the sellar region** | | | | n/a | | |
| Tumors of the pituitary gland | 8140/0 | Adenoma, NOS | C75.1 | n/a | n/a | 3,995 |
| | 8140/1 | Atypical adenoma<br>*Synonyms:* Bronchial adenoma, NOS | C75.1 | n/a | n/a | <16 |

| | | | | | | |
|---|---|---|---|---|---|---|
| | 8140/3 | Adenocarcinoma, NOS<br>*Synonyms:* Adenocarcinoma, usual type | C75.1 | n/a | n/a | <16 |
| | 8146/0 | Monomorphic adenoma | C75.1 | n/a | n/a | <16 |
| | 8246/3 | Neuroendocrine carcinoma, NOS<br>*Synonyms:* Poorly differentiated neuroendocrine neoplasm | C75.1 | n/a | n/a | <16 |
| | 8260/0 | Papillary adenoma, NOS | C75.1 | n/a | n/a | 17 |
| | 8270/0 | Chromophobe adenoma | C75.1 | n/a | n/a | 124 |
| | 8270/3 | Chromophobe carcinoma<br>*Synonyms:* Chromophobe adenocarcinoma | C75.1 | n/a | n/a | <16 |
| | 8271/0 | Lactotroph adenoma<br>*Synonyms:* Prolactinoma | C75.1, C75.2 | n/a | n/a | 4,059 |
| | 8272/0 | Pituitary adenoma, NOS | C75.1, C75.2 | n/a | n/a | 76,517 |
| | 8272/3 | Pituitary carcinoma, NOS | C75.1 | n/a | n/a | 438 |
| | 8280/0 | Acidophil adenoma<br>*Synonyms:* Eosinophil adenoma | C75.1 | n/a | n/a | 21 |
| | 8281/0 | Mixed acidophil-basophil adenoma | C75.1 | n/a | n/a | <16 |
| | 8290/0 | Oxyphilic adenoma<br>*Synonyms:* Oncocytic adenoma; Oncocytoma | C75.1 | n/a | n/a | 40 |
| | 8300/0 | Basophil adenoma<br>*Synonyms:* Mucoid cell adenoma | C75.1 | n/a | n/a | <16 |
| | 8320/3 | Granular cell carcinoma<br>*Synonyms:* Granular cell adenocarcinoma | C75.1 | n/a | n/a | <16 |
| | 8323/0 | Mixed cell adenoma | C75.1 | n/a | n/a | <16 |
| | 9432/1 | Pituicytoma | C75.1 | n/a | n/a | 67 |
| | 9580/0 | Granular cell tumor, NOS<br>*Synonyms:* Granular cell myoblastoma, NOS | C75.1, C71.5 | n/a | n/a | <16 |
| | 9580/3 | Granular cell tumor, malignant<br>*Synonyms:* Granular cell myoblastoma, malignant | C75.1 | n/a | n/a | <16 |
| | 9582/0 | Granular cell tumor of sellar region | C75.1, C71.9 | n/a | n/a | 29 |
| Craniopharyngioma | 9350/1 | Craniopharyngioma, NOS<br>*Synonyms:* Rathke pouch tumor | C70-C72, C75.1- C75.3 | n/a | n/a | 1,303 |
| | 9351/1 | Craniopharyngioma, adamantinomatous | C70-C72, C75.1- C75.3 | n/a | n/a | 1,479 |
| | 9352/1 | Craniopharyngioma, papillary | C70-C72, C75.1- C75.3 | n/a | n/a | 372 |
| **Neoplasm, unspecified** | 8000/0 | Neoplasm, benign<br>*Synonyms:* Tumor, benign; Unclassified tumor, benign | C70-C72, C75.1- C75.3 | n/a | n/a | 3,332 |
| | 8000/1 | Neoplasm, uncertain whether benign or malignant<br>*Synonyms:* Neoplasm, NOS; Tumor, NOS; Unclassified tumor, borderline malignancy; Unclassified tumor, uncertain whether benign or malignant | C70-C72, C75.1- C75.3 | n/a | n/a | 3,264 |
| | 8000/3 | Neoplasm, malignant<br>*Synonyms:* Blastoma, NOS; Tumor, malignant, NOS; Cancer; Malignancy; Unclassified tumor, malignant | C70-C72, C75.1- C75.3 | n/a | n/a | 7,274 |
| | 8001/0 | Tumor cells, benign | C70-C72, C75.1- C75.3 | n/a | n/a | 37 |

| | | | | | | |
|---|---|---|---|---|---|---|
| | 8001/1 | Tumor cells, uncertain whether benign or malignant<br>*Synonyms:* Tumor cells, NOS | C70-C72, C75.1- C75.3 | n/a | n/a | 30 |
| | 8001/3 | Tumor cells, malignant | C70-C72, C75.1- C75.3 | n/a | n/a | 20 |
| | 8002/3 | Malignant tumor, small cell type | C70-C72, C75.1- C75.3 | n/a | n/a | <16 |
| | 8003/3 | Malignant tumor, giant cell type | C70-C72, C75.1- C75.3 | n/a | n/a | <16 |
| | 8004/3 | Malignant tumor, spindle cell type<br>*Synonyms:* Malignant tumor, fusiform cell type | C70-C72, C75.1- C75.3 | n/a | n/a | 17 |
| | 8005/0 | Clear cell tumor, NOS | C70-C72, C75.1- C75.3 | n/a | n/a | <16 |
| | 8005/3 | Malignant tumor, clear cell type | C70-C72, C75.1- C75.3 | n/a | n/a | <16 |
| | 8010/0 | Epithelial tumor, benign | C70-C72, C75.1- C75.3 | n/a | n/a | <16 |
| | 8010/3 | Carcinoma, NOS<br>*Synonyms:* Epithelial tumor, malignant | C70-C72, C75.1- C75.3 | n/a | n/a | 24 |
| **All other** | 8713/0 | Glomangiomyoma | C70-C72, C75.1- C75.3 | n/a | n/a | <16 |
| | 9173/0 | Cystic lymphangioma<br>*Synonyms:* Hygroma, NOS; Cystic hygroma | C70-C72, C75.1- C75.3 | n/a | n/a | 63 |
| | 9503/3 | Neuroepithelioma, NOS | C70-C72, C75.1- C75.3 | n/a | n/a | 57 |

**Supplementary Figure 1. Overview of recode process for specific tumor types**

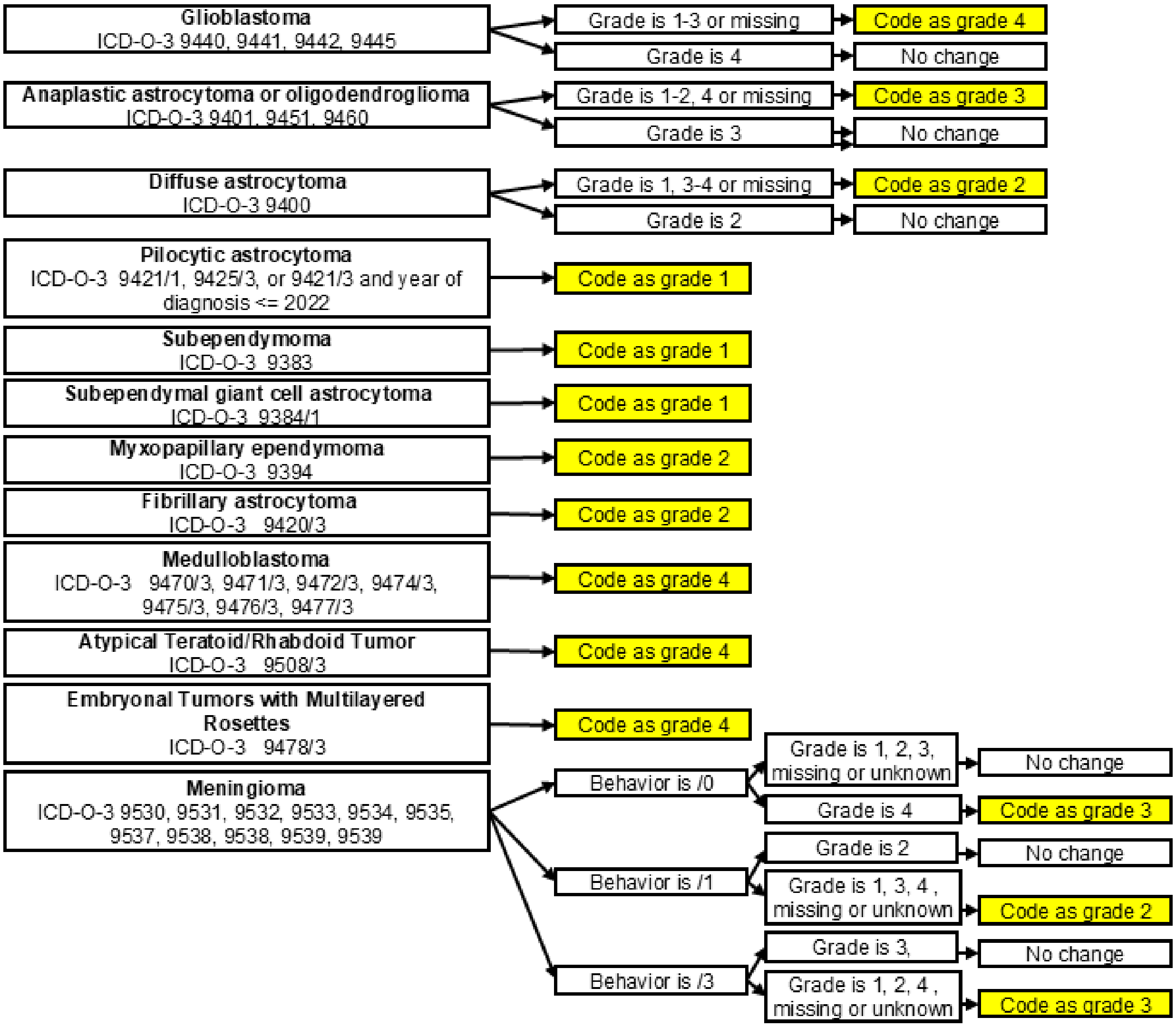

**Supplementary Figure 2. A) Grade of glioma cases before and after grade recoding process, B) distribution of consensus grade in glioma cases, C) distribution of grade in glioma cases post-recoding process, and D) overall survival for IDHmut astrocytoma under the consensus and recoded grading schemes**

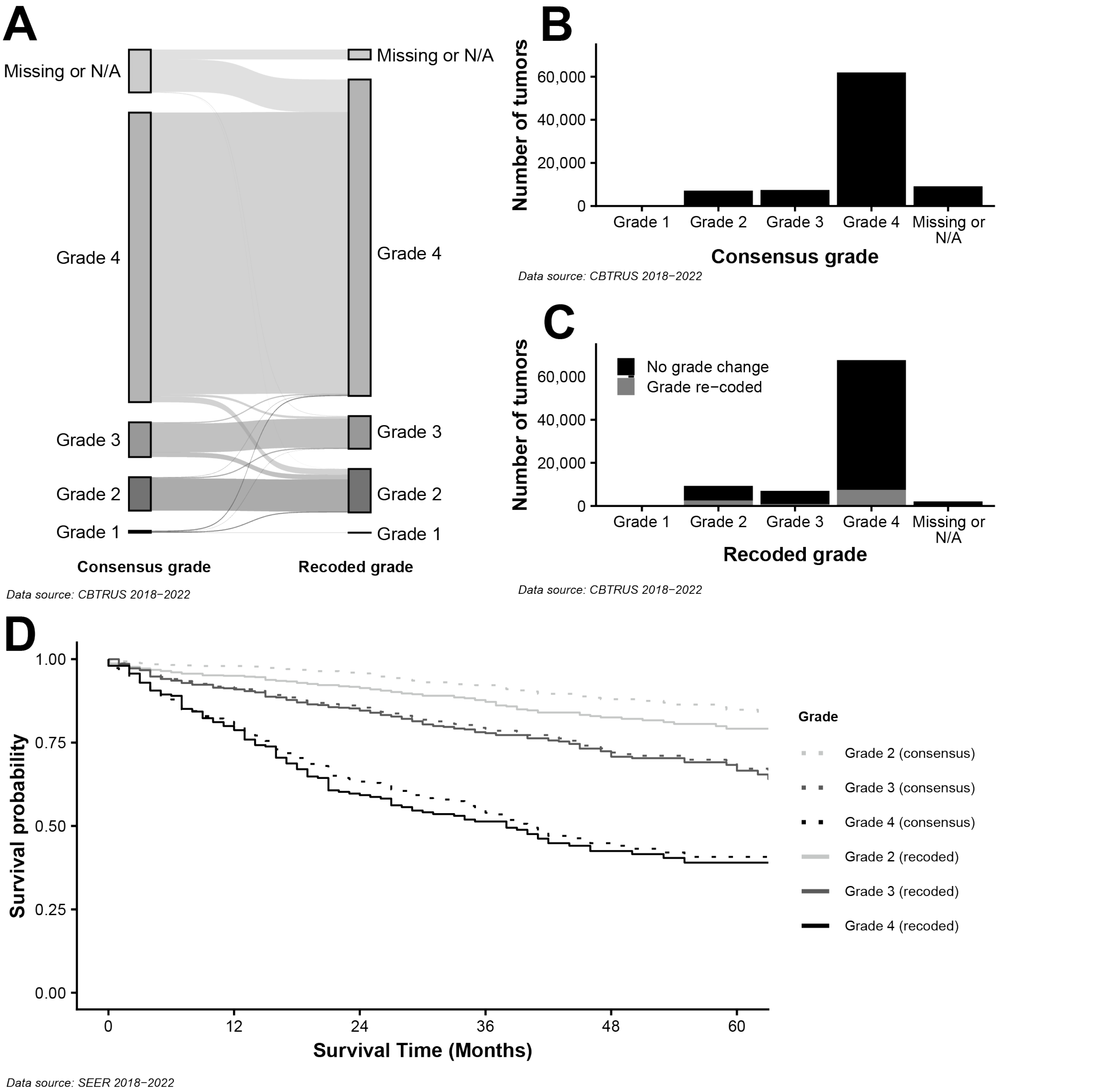

**Supplementary Figure 3. Completeness of the Brain Molecular Marker Variable by ICD-O-3 Code and Year of Diagnosis**

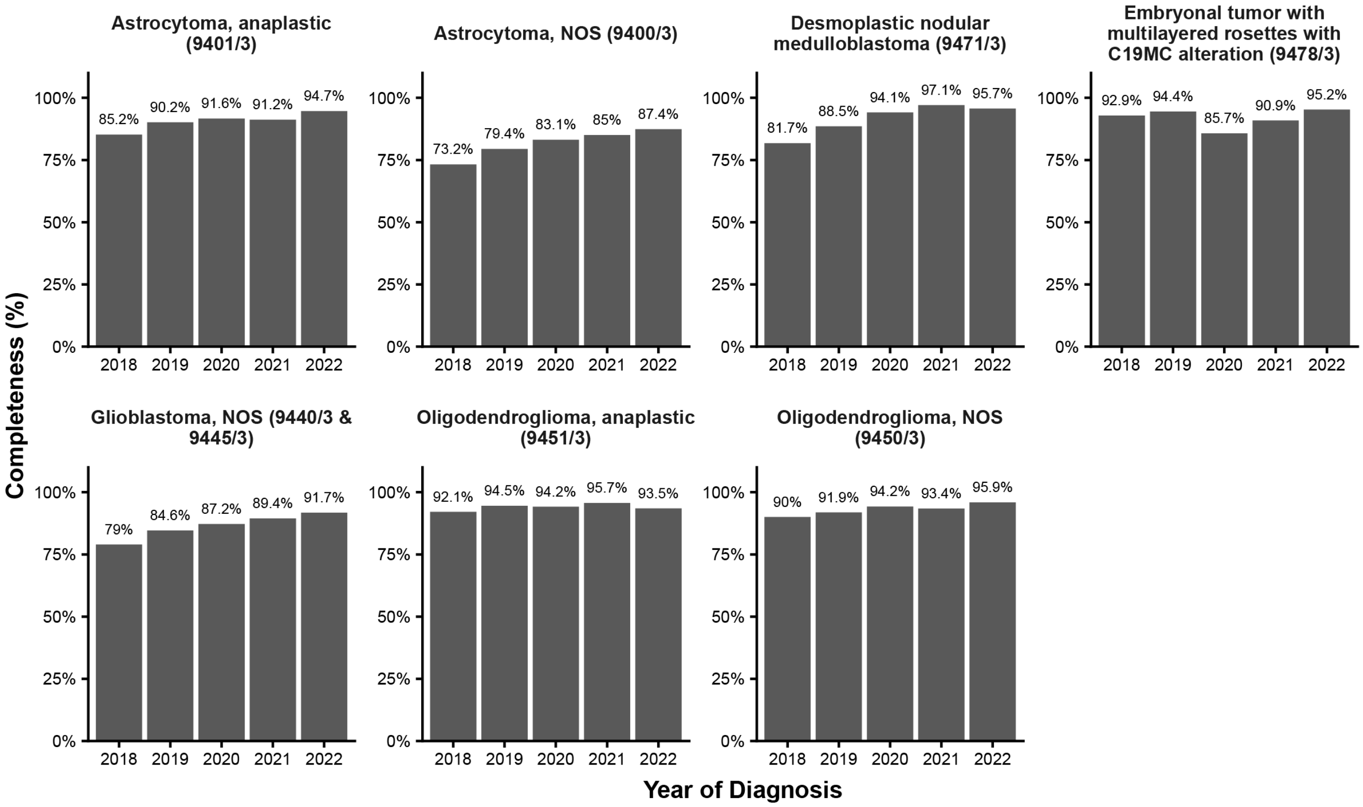

*Data source: CBTRUS 2018−2022*

**Supplementary Figure 4. Changes in case classification between 2021 and 2026 scheme for a) selected glioma entities, B) CNS lymphoma, and C) other tumor types**

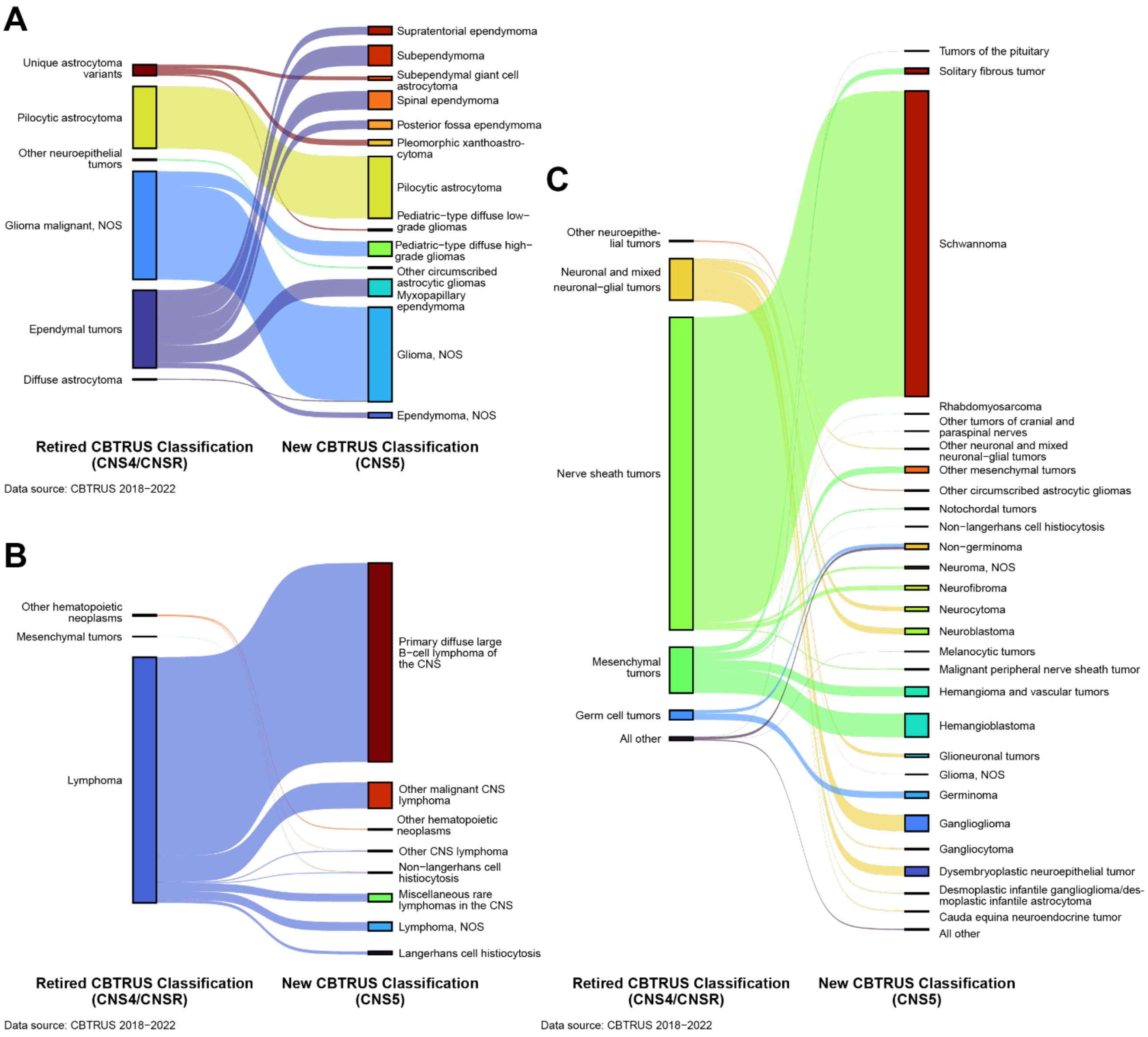
